# Genetic and Causal Associations Between Tobacco Smoking and Mental Health After Accounting for General Substance Use and Socioeconomic Factors

**DOI:** 10.64898/2026.07.22.26358653

**Authors:** Adrià Túnez, Dirk J.A. Smit, Abdel Abdellaoui, Anil P.S. Ori, Jorien L. Treur, Joëlle A. Pasman, Karin J.H. Verweij, 23andMe Research Team

**Affiliations:** Amsterdam UMC location University of Amsterdam, Department of Psychiatry, Meibergdreef 9, Amsterdam, the Netherlands; Amsterdam Public Health Research Institute, Amsterdam, The Netherlands; Amsterdam Neuroscience Research Institute, Amsterdam, The Netherlands; 23andMe Research Institute, 2555 Park Blvd Palo Alto, CA 94306 USA

## Abstract

Genetic instrumental variable studies can provide stronger insights into the causal relationships between smoking and mental illness than conventional observational studies because they are less susceptible to confounding and reverse causation. However, they still rely on genetic instruments that partly capture genetic influences shared with other substance use and socioeconomic status (SES), potentially biasing estimates of smoking-specific effects. We therefore aim to (1) develop a more specific genetic instrument for smoking that minimizes these shared influences and (2) use this instrument to examine the causal effects of smoking on psychiatric disorders.

We applied Genomic Structural Equation Modelling to 19 European-ancestry GWAS summary statistics (7 smoking, 8 substance use and 4 SES phenotypes), deriving a novel smoking-specific genetic factor representing liability to smoking independent of shared substance-use and SES influences. We used this factor as an instrument in Mendelian Randomization analyses to test causal effects of smoking on eight psychiatric disorders.

The smoking-specific factor was associated with 52 independent genome-wide significant loci. Genetically predicted smoking-specific liability was significantly causally associated with seven psychiatric disorders. These findings support a causal role of smoking in increasing the risk of multiple psychiatric disorders beyond influences shared with other substance use and SES, providing stronger evidence for smoking-specific effects and informing targeted smoking prevention and intervention strategies. More broadly, our findings highlight that the validity of Mendelian Randomization depends on the specificity of its genetic instruments. Future studies should strive to develop instruments that better isolate the exposure of interest from shared genetic influences, enabling more accurate identification of causal mechanisms.

## Introduction

Smoking is one of the most widespread preventable risk factors for poor health globally, contributing significantly to disease burden (1). Beyond its role in cancer, cardiovascular, and respiratory diseases, smoking is comorbid with mental illnesses (2). For instance, in 2019 in the US, 33% of people with a serious mental illness were smokers compared to 16% of the general population (3). A 2021 factsheet by the WHO revealed that 2 in 3 people with severe mental illness were smokers (4). Among people with schizophrenia, smoking prevalence even reaches 60% (5, 6), substantially contributing to the already elevated morbidity and mortality rates in these populations.

It has long been assumed that the association between mental illness and smoking is the result of (symptoms of) mental illness increasing the likelihood of smoking as a maladaptive coping mechanism, or due to impaired decision-making (7). However, there is now converging evidence from multiple research methods that there are also causal effects in the opposite direction: smoking increasing the risk or severity of mental illness (8–13). While these findings are consistent across several methodological approaches, the evidence base has limitations - including potential residual confounding, and challenges in establishing temporality - and should be interpreted with appropriate caution. Nevertheless, these insights have important public health implications, reshaping health messaging and prevention strategies.

Since randomized controlled trials on smoking are largely unfeasible, advances in genetics have played a key role in establishing this causal association. Genome-wide association studies (GWAS) have identified many thousands of genetic variants associated with individual differences in a trait of interest (14). Mendelian Randomization (MR) leverages these genetic variants associated with exposure traits - such as smoking behaviours - as instrumental variables to test causal effects on outcomes (15). MR studies using GWAS summary statistics have reported evidence suggesting that smoking increases the risk of psychiatric disorders, including MDD and schizophrenia (16, 17).

Two major limitations hinder the interpretability and precision of current MR studies on smoking and mental health. First, genetic associations identified by GWASs do not only capture trait-specific signal; signals are often biased by confounding or pleiotropic traits and gene-environment correlations (18). In the case of smoking, it is highly likely that the SNP associations for smoking are confounded by use of other substances and socioeconomic status (SES) (19–24). For instance, studies show that individuals with lower SES are more likely to be exposed to tobacco smoke, start smoking earlier, and smoke more heavily (25–27). GWAS designs typically do not model or remove such confounding, meaning the resulting genetic associations may not purely reflect smoking-specific liability. As a result, SNPs derived from these GWASs may not be valid instruments for smoking in MR analyses, as they risk conflating smoking-specific effects with those of correlated traits such as broader substance use or socioeconomic disadvantage. This is especially relevant in MR studies, where SNPs for which there is no knowledge of a biological function are often included, increasing the chance of bias (28). Recent studies have aimed at disentangling the shared genetic architecture of substance use traits (29, 30), but none have specifically isolated the genetic signal unique to smoking from that of its most important correlated confounders such as broader substance use and SES.

Second, the genetic instruments currently used in MR to capture smoking behaviour are derived from GWASs that used heterogeneous phenotyping strategies across studies: depending on which GWAS is used, an instrument may reflect cigarettes per day, age at smoking initiation, or another single facet of tobacco use, and these facets are typically derived from self-reported questionnaire items or meta-analyses that merge cohorts with non-aligned definitions (31). This instrument-level heterogeneity is problematic for interpretation, since the causal estimates obtained can depend on which smoking GWAS is chosen as the source of instruments. At the same time, this proliferation of GWASs reflects genuine progress: in recent years, several GWASs have provided more measures of smoking phenotypes, including smoking initiation, cessation, age of onset, tobacco use disorder, and nicotine dependence (32–34). These studies have helped characterize different dimensions of the genetic architecture underlying smoking and identified a large number of associated genetic variants. Nevertheless, a substantial gap remains: although twin studies estimate the heritability of smoking behaviours at 40–80% (35, 36), SNP-based heritability estimates currently account for only around 25% of this variance (32, 37). In 2022, Wootton et al. conducted a GWAS of lifetime smoking behaviour, which captured smoking duration, heaviness, and cessation (31). However, a comprehensive GWAS that integrates all the available facets of smoking behaviour is still lacking, while such an approach may be crucial for substantially increasing power and capturing the broader genetic liability to tobacco use.

Shedding light on the genetic signal that is unique to smoking, and not shared with these important confounders, while also leveraging the shared signal across smoking phenotypes, would help identify more precise instruments to disentangle whether there are causal relationship between tobacco smoking and mental health problems. This would also clarify whether previously observed causal associations in MR studies could be explained by any confounding factors that are captured by GWASs.

To overcome these limitations, our study pursued two goals. First, we applied Genomic Structural Equation Modelling (Genomic SEM) (38) to combine existing tobacco related GWASs and disentangle shared and unique sources of genetic variance among smoking, general substance use, and SES-related traits. This enabled us to extract a latent factor reflecting smoking-specific genetic liability. Second, we used the smoking-specific factor to examine the causal relationship between smoking and eight major psychiatric disorders using Mendelian Randomization. By reducing bias and improving specificity in our exposure variable, we aimed to produce more robust and interpretable causal estimates.

Together, these efforts address two gaps in the field: the need for more precise genetic instruments, and the need for clearer evidence on the causal role of smoking in mental health. Ultimately, this work may inform more targeted public health interventions and improve risk communication strategies around tobacco use and mental illness.

## Results

Full methods and results for the Townsend Deprivation Index and Caffeine Use GWASs can be found in the Supplementary Information.

### 1. Latent factor architecture mapping

To assess how genetically similar the model traits were, we calculated SNP-based heritability estimates and genetic correlations between all nineteen traits (Figure 2; GWAS sources in Supplementary Table 1). All correlations were in the expected direction, and most traits showed moderate to high genetic correlations. In particular, all tobacco-related traits showed moderate to high correlations (absolute correlations ranging from 0.31 to 0.9; all p<.05 – FDR corrected) as well as all SES related traits (0.55 to 0.91; all p<.05). The other substance use traits were more heterogeneous: correlations were generally high between different traits indexing the same substance, but more modest to high across different substances, and across substance use traits and SES or smoking traits. The caffeine and alcohol use traits showed the weakest correlations with other traits. Smoking related traits also strongly overlapped with both substance use and SES-related traits.

**Figure 1.**
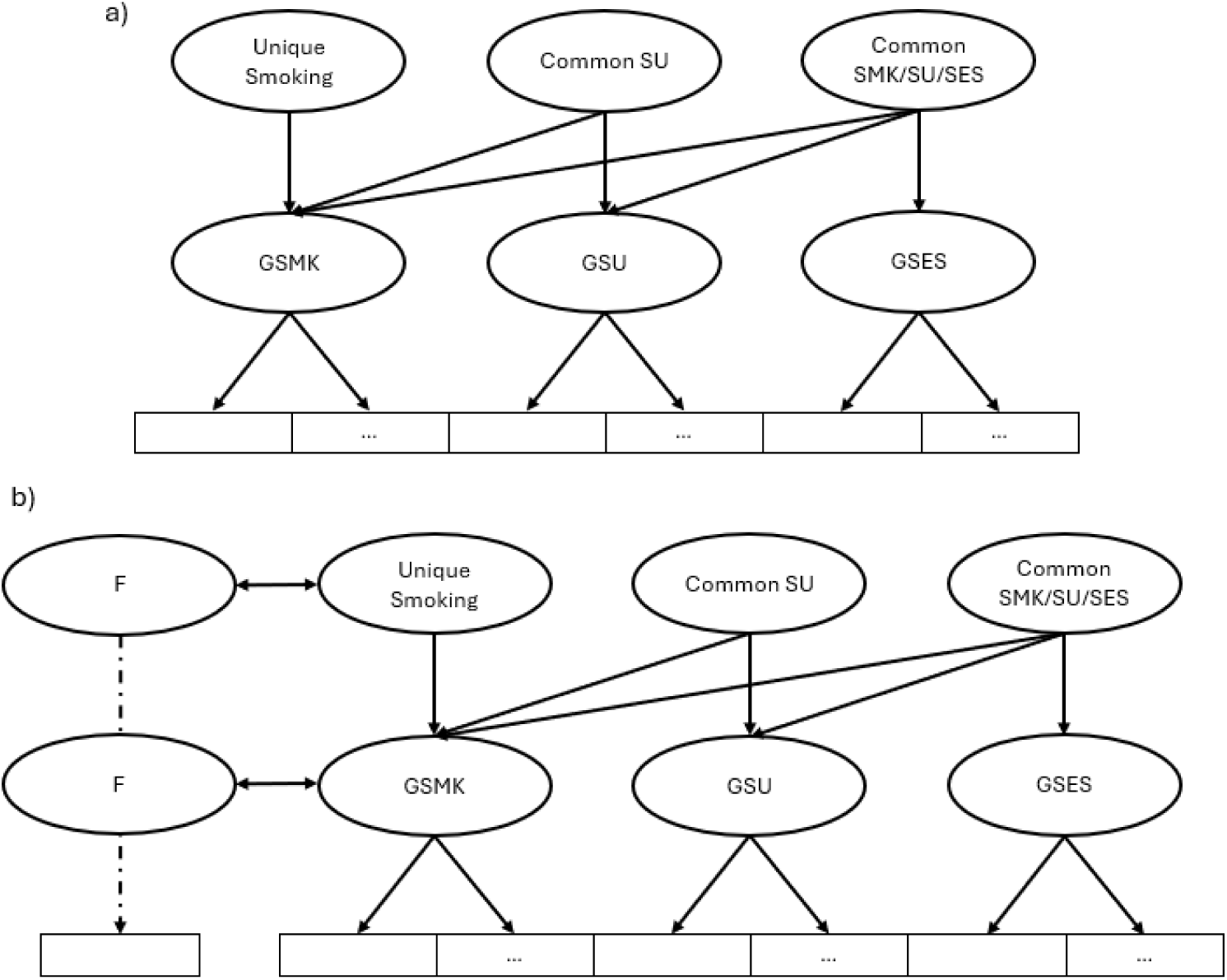
**a)** Schematic representation of the two-layered hierarchical Genomic SEM model. Observed traits (which correspond to GWAS summary statistics) are represented as squares, latent factors as circles. Given the large number of traits used, they are not mentioned in this schematic figure. **b)** Schematic representation of the two-layer hierarchical Genomic SEM model including genetic correlations with other factors. An observed trait is first modelled as a latent factor (F). In the first level, F is allowed to correlate with GSMK, in the second level, it is allowed to correlate with the *Unique Smoking Factor*. GSMK = General Smoking Factor, GSU = General Substance Use Factor, GSES = General Socioeconomic Status Factor, F = Factor.

**Figure 2.**
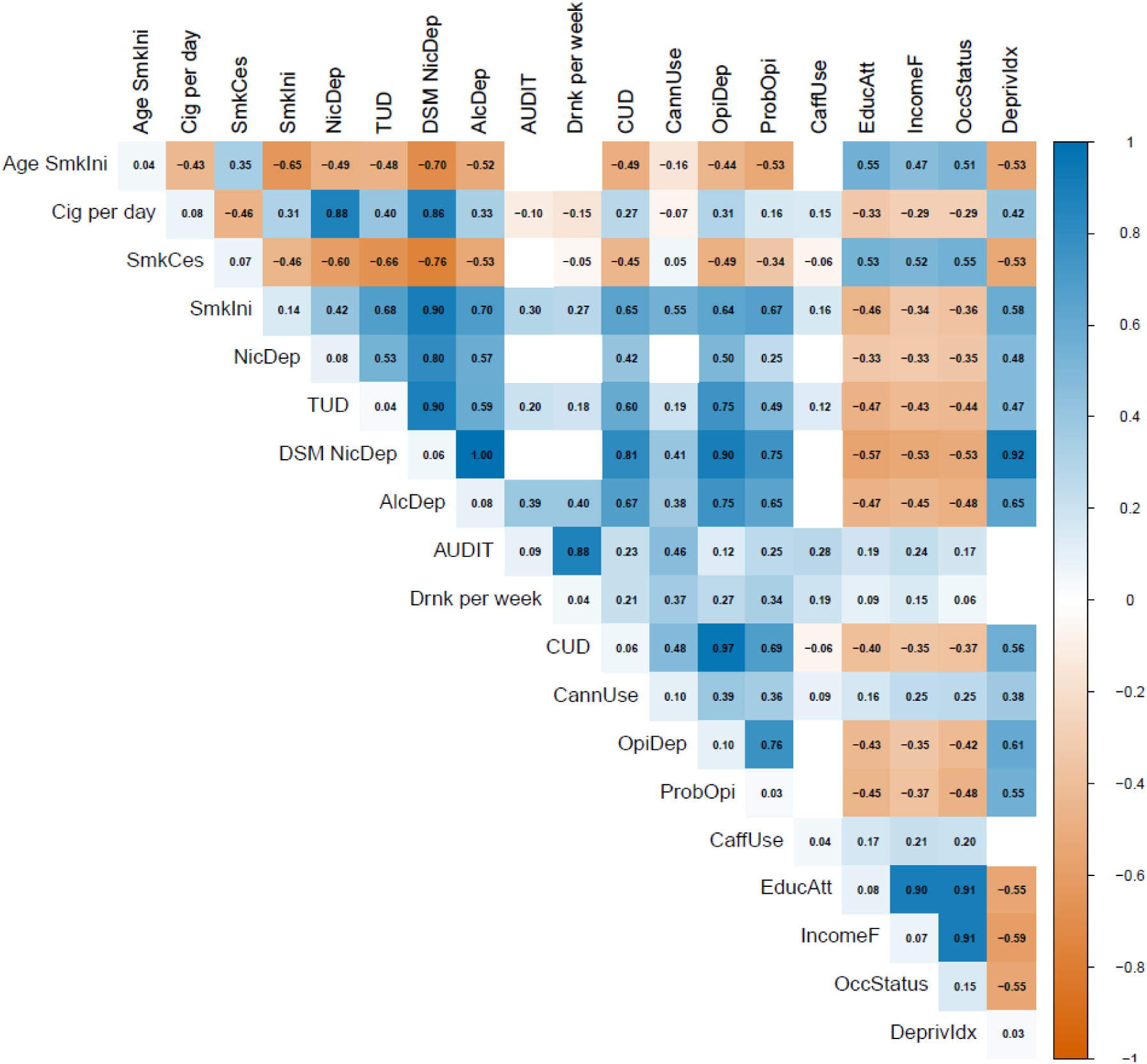
Genetic correlation heatmap of all 19 GWAS summary statistics used for the Genomic SEM model. Numbers represent genetic correlations values. Positive correlations are depicted in blue, negative in orange. Darker colours indicate stronger correlations. Diagonal cells show SNP−based heritability (h2) on the liability scale. Blank off−diagonal cells indicate genetic correlations not significant after FDR multiple testing correction (p < .05). Acronyms for trait names are detailed in the summary statistics table (Supplementary Table 1).

The objective of our proposed model was to tease apart genetic signal that is unique to smoking behaviours from signal shared with other substance use or SES traits. Given the low genetic correlations of caffeine use with other traits and its non-significant loading onto GSU, it was excluded from the final model. Our model, in which we first model three general latent factors (GSMK, GSU, GSES) and then model three hierarchical latent factors (*Unique Smoking*, Common SU, Common SMK/SU/SES), fit the data well (CFI=0.9, SRMR=0.15; close to the standard thresholds of good fit CFI=0.9 SRMR=0.1). Factor loadings are shown in Figure 3 (full results in Supplementary Table 2). Of special interest are the 0.50 loadings of the common SU factor on GSMK p<0.05, and the −0.72 loadings of the common SMK/SU/SES factor on GSMK p<0.05, which show that part of the general smoking genetic signal overlaps with the genetic signal of general substance use and general SES respectively.

**Figure 3.**
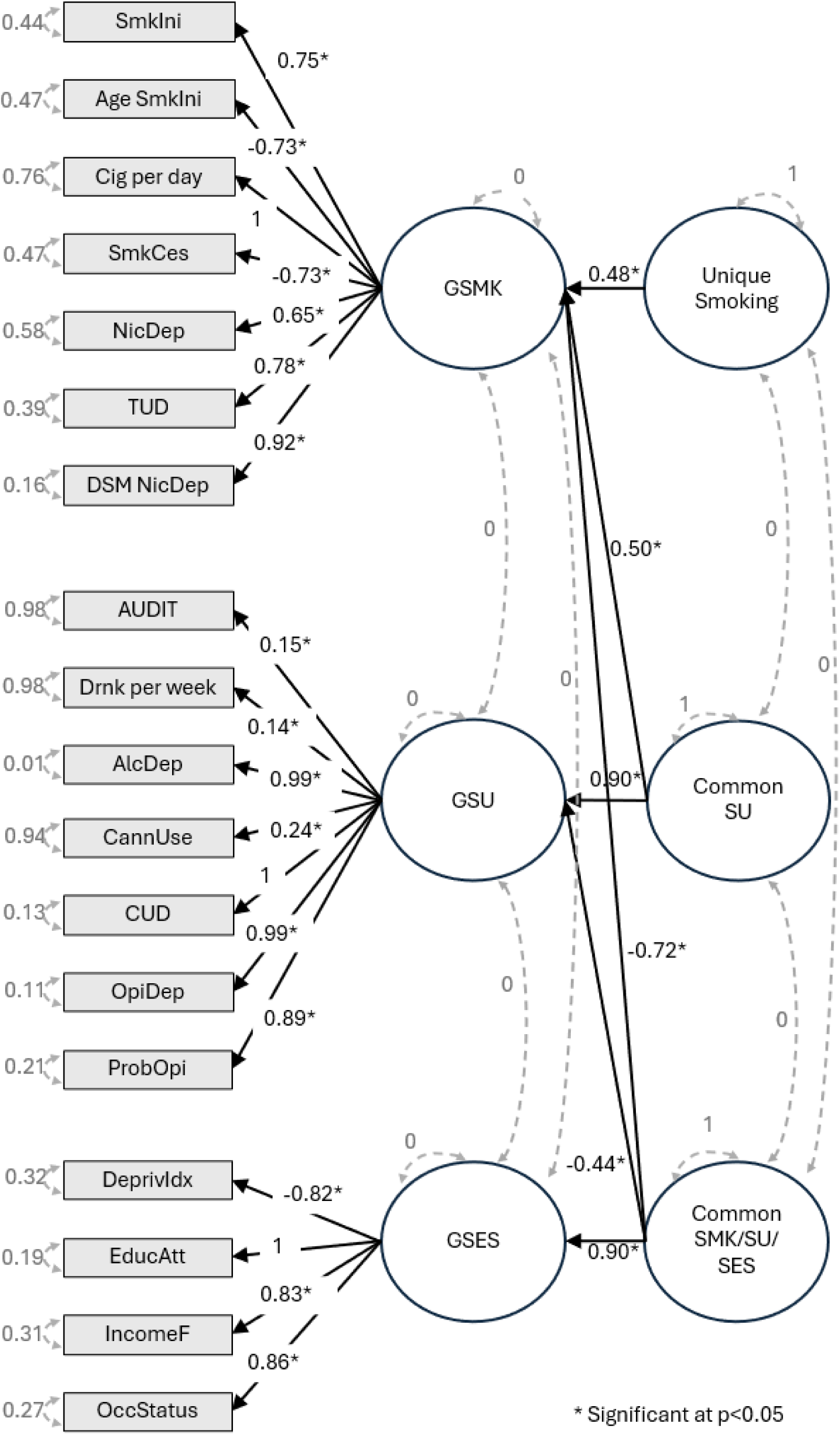
Genomic SEM model results. Standardized factor loadings are represented in black text and black arrows. Residual variances and between-factor correlations are represented in grey text and grey dotted lines. Circles represent latent factors, and rectangles are “observed” variables (Summary-level GWAS data). Asterisks represent significance at nominal level p<0.05. GSMK, general smoking liability; GSU, general substance use liability; GSES, general SES liability; Common SU, common substance use; Common SMK/SU/SES, common smoking, substance and SES.

The 0.48 loading of the *Unique Smoking Factor* on GSMK p<0.05 indicates that there is a genetic component of smoking that is not shared with either other substance use or socioeconomic traits.

### 2. Evaluation of the Unique Smoking Factor

Under our model specifications, we compared genetic correlations of GSMK and the *Unique Smoking Factor* with the smoking, substance use, and SES traits in the model to test whether removing shared genetic signal with substance use and SES produced the expected patterns. For smoking traits with higher overlap with substance use/SES in the original LDSC correlations (age of smoking initiation, smoking initiation, cessation, DSM nicotine dependence, tobacco use disorder), correlations with the *Unique Smoking Factor* remained similar to GSMK (albeit significantly different for cessation and tobacco use disorder), while traits with lower overlap (cigarettes per day, nicotine dependence) showed significantly higher correlations with the *Unique Smoking Factor* than with GSMK — in line with expectations. For other substance use traits, all correlations with the *Unique Smoking Factor* were non-significant or decreased (except for drinks per week), compared to significant positive correlations with GSMK, again as expected. Among SES traits, educational attainment’s correlation became non-significant with the *Unique Smoking Factor*, all correlations of SES traits with the *Unique Smoking Factor* decreased as expected. However, income and occupational status showed a significant sign reversal — from negative with GSMK to positive with the *Unique Smoking Factor* (Figure 4).

**Figure 4.**
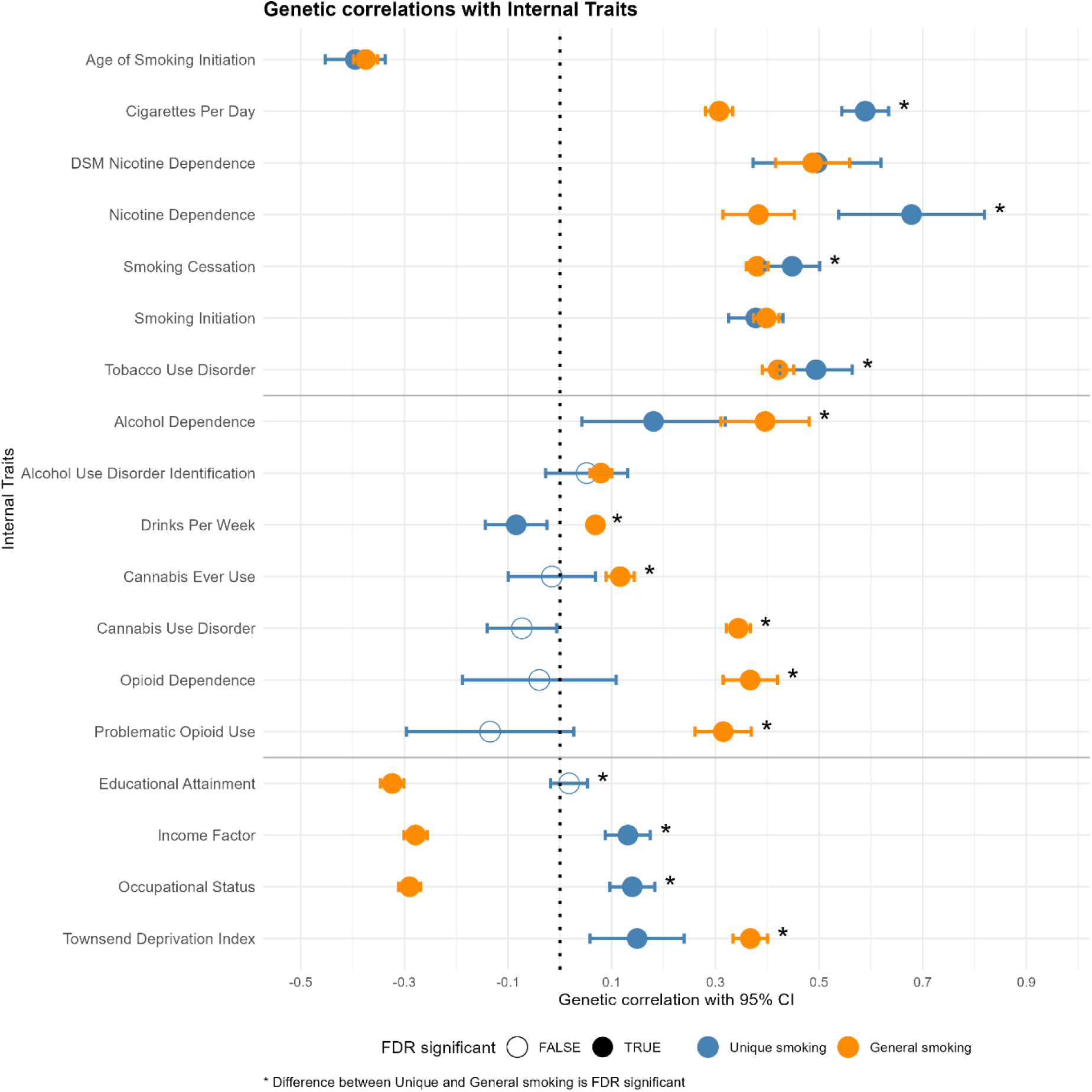
Forest plot depicting the genetic correlations of the general (GSMK-in orange) and unique (*Unique Smoking Factor*-in blue) with all substance use and SES traits . Empty dots represent non-significant correlations. Filled dots represent significant genetic correlations after FDR multiple testing correction. Stars indicate significant difference of the genetic correlations between GSMK and unique smoking factors after multiple testing correction using the delta method. CI, confidence interval.

GWAS on the *Unique Smoking Factor* revealed 62 lead SNPs (r² < 0.1) across 52 genomic risk loci. Note that the signal was inflated, lambda = 1.37, a finding that is common in GenomicSEM due to signal boost from aggregation of shared heritable variance across traits, and scaling of residual variance (39). Nonetheless, the LDSC intercept = 0.93 suggests the inflation is not due to population stratification. Among the strongest hits were SNPs in the *CHRNB4, CHRNA3,* and *CHRNA5* nicotine receptor genes, and *CADM2* which are also significant hits in the individual smoking GWASs. The Manhattan plot is depicted in Figure 5a. Gene-based test identified 181 significant genes, among which were the nicotinic receptor genes, as well as genes involved in the effects of nicotine in dopaminergic pathways (Supplementary Figure 1), Tissue expression analyses showed enrichment in several brain tissues, with the strongest signals observed in the frontal cortex, anterior cingulate cortex, nucleus accumbens, cerebellum, and basal ganglia regions (Figure 5b).

**Figure 5.**
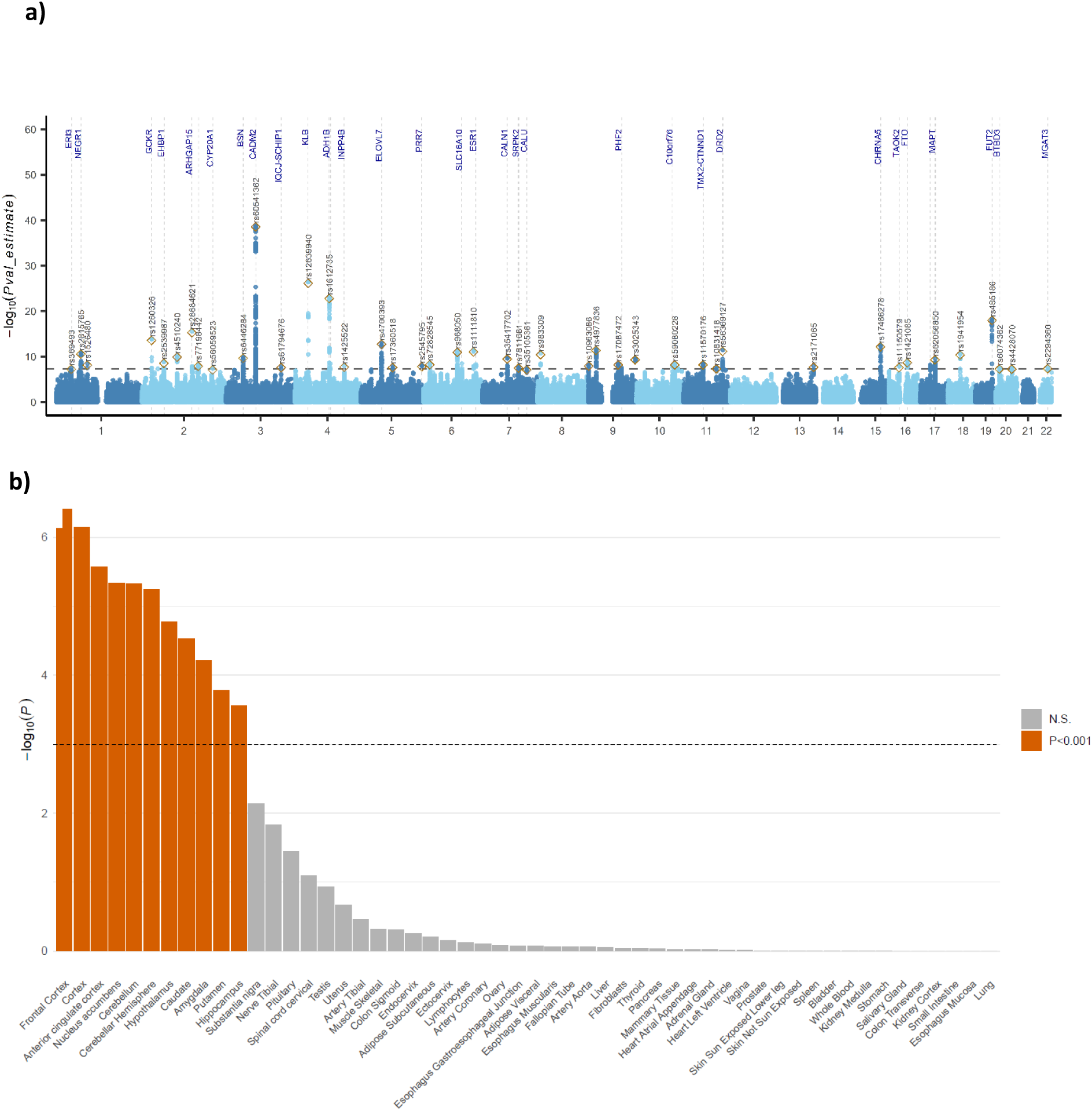
**a)** Manhattan plot of the latent *Unique Smoking Factor* GWAS. The red line indicates the genome-wide significance threshold of p <5e−8. **b)** Tissue expression enrichment results from FUMA using GTEx v8 data for genes associated with the *Unique Smoking Factor*. The dashed horizontal line indicates the significance threshold after multiple-testing correction.

### 3. Genetic correlations between GSMK and the Unique Smoking Factor with mental health indices

To better understand the association between smoking and mental health, we first evaluated the genetic association between the *Unique Smoking Factor* and eight common mental health outcomes (Figure 6). All psychiatric traits except OCD showed a significant genetic correlation with GSMK, in line with previous literature linking smoking to mental health. With the *Unique Smoking Factor*, however, only MDD, OCD, bipolar, and PTSD remained significantly correlated, albeit this difference was not FDR-significant in bipolar disorder. For anorexia nervosa, anxiety disorders, insomnia, the correlation with the *Unique Smoking Factor* was attenuated to non-significance, while retaining the same direction as GSMK; this difference was not FDR-significant in anorexia nervosa. MDD and PTSD remained significantly and positively correlated with the *Unique Smoking Factor*, though with smaller effect sizes than GSMK. OCD showed a notable pattern: its correlation with the *Unique Smoking Factor* was significant and negative, in contrast to a non-significant correlation with GSMK — a significant reversal that stands out from the broader pattern of attenuation. This pattern, whereby the genetic association between GSMK and several psychiatric outcomes is substantially reduced once shared variance with substance use and SES is removed, suggests that part of this association may be attributable to confounding rather than smoking-specific liability. This is further considered in the Discussion in relation to the conditional nature of the latent factors.

**Figure 6.**
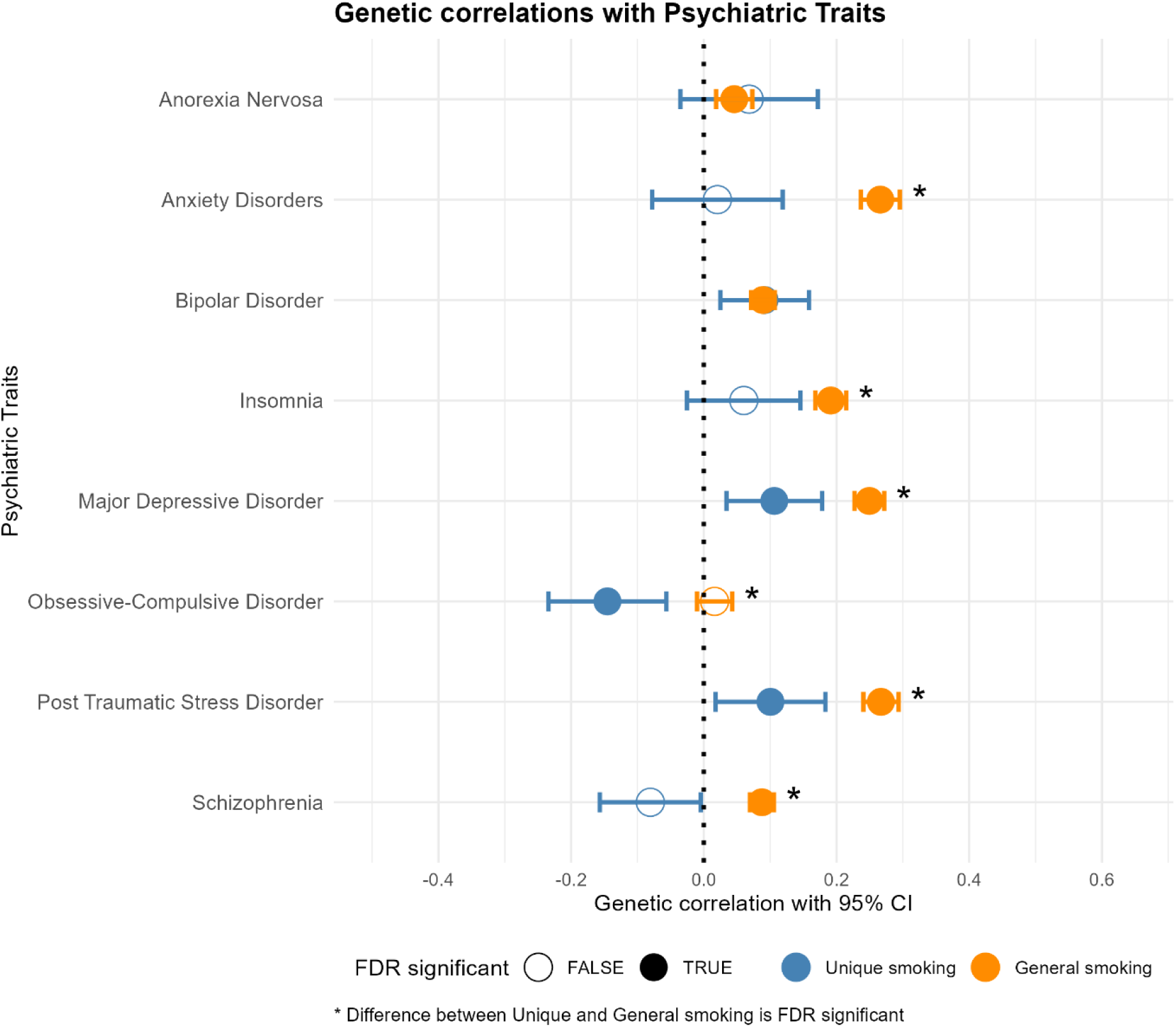
Forest plot depicting the genetic correlations with 95% CIs of the general (GSMK-in orange) and unique (*Unique Smoking Factor* -in blue) smoking factors with eight common psychiatric traits. Filled dots represent significant genetic correlations after FDR multiple testing correction. Stars represent significant differences between the general and unique smoking factors after multiple testing correction using the delta method. CI, confidence interval.

### 2.4. Examining causal associations of smoking with mental health through MR

We then obtained instrumental variables from the *Unique Smoking Factor* to test causal associations between smoking and eight mental health outcomes. We incorporated individual SNP effects in the Genomic SEM model to perform Mendelian Randomization analysis to test causal associations of smoking on mental health outcomes while modelling potential confounders. While this does not guarantee complete independence from all confounders, it allows us to explicitly model and reduce bias from substance use and SES. The 56 genome-wide significant SNPs used as instruments explained a substantial proportion of variance in the *Unique Smoking Factor* (mean per-SNP R² = 0.002; total R² = 0.12; mean F-statistic = 43.9). Importantly, this R² reflects variance explained on the latent-factor scale derived from Genomic SEM, rather than phenotypic variance in an observed smoking measure. These results are not directly comparable to those obtained from conventional single-trait GWAS as used in general two-sample MR. As such, the elevated total R² likely reflects a combination of multivariate signal aggregation and scaling inherent to the Genomic SEM framework, rather than unusually strong phenotypic instruments. Results showed significant (*p* < 0.05) associations in line with *Unique Smoking* causally increasing the risk of all common psychiatric trait liabilities except OCD. The largest effects were observed for anxiety disorder, MDD, and PTSD liability (unstandardized estimates = 0.21-0.24). OCD and *Unique Smoking* were not significantly associated. Among the 56 instrumental SNPs used as proxy for *Unique Smoking*, several showed significant direct effects on the outcomes, thus affecting those via paths distinct than the exposure, which could compromise the exclusion restriction assumption. MDD showed 16 pleiotropic SNPs, bipolar disorder 10, schizophrenia 11, anorexia nervosa 5, anxiety 7, PTSD 9, and insomnia 4. This finding shows that there are biological pathways distinct from smoking through which these SNPs exert their effects on psychiatric outcomes. By independently accounting for these pleiotropic SNPs in the Genomic SEM framework, we aimed to account for violations of the MR exclusion restriction assumption and reduce bias in causal estimates. After accounting for pleiotropic effects, the causal effect estimates remained robust. We conclude that these results indicate that there is a causal effect from smoking on all psychiatric traits except OCD. All MR results are depicted in Figure 7a. Out of 56 SNPs for the *Unique Smoking Factor* that had a significant effect on the psychiatric traits, 26 were shared across all traits and 20 were shared among at least 6 out of the 7 outcomes significantly affected by smoking. The remaining were shared among 5 or fewer outcomes. Visual representation of the SNP overlap can be seen in Figure 7b.

**Figure 7.**
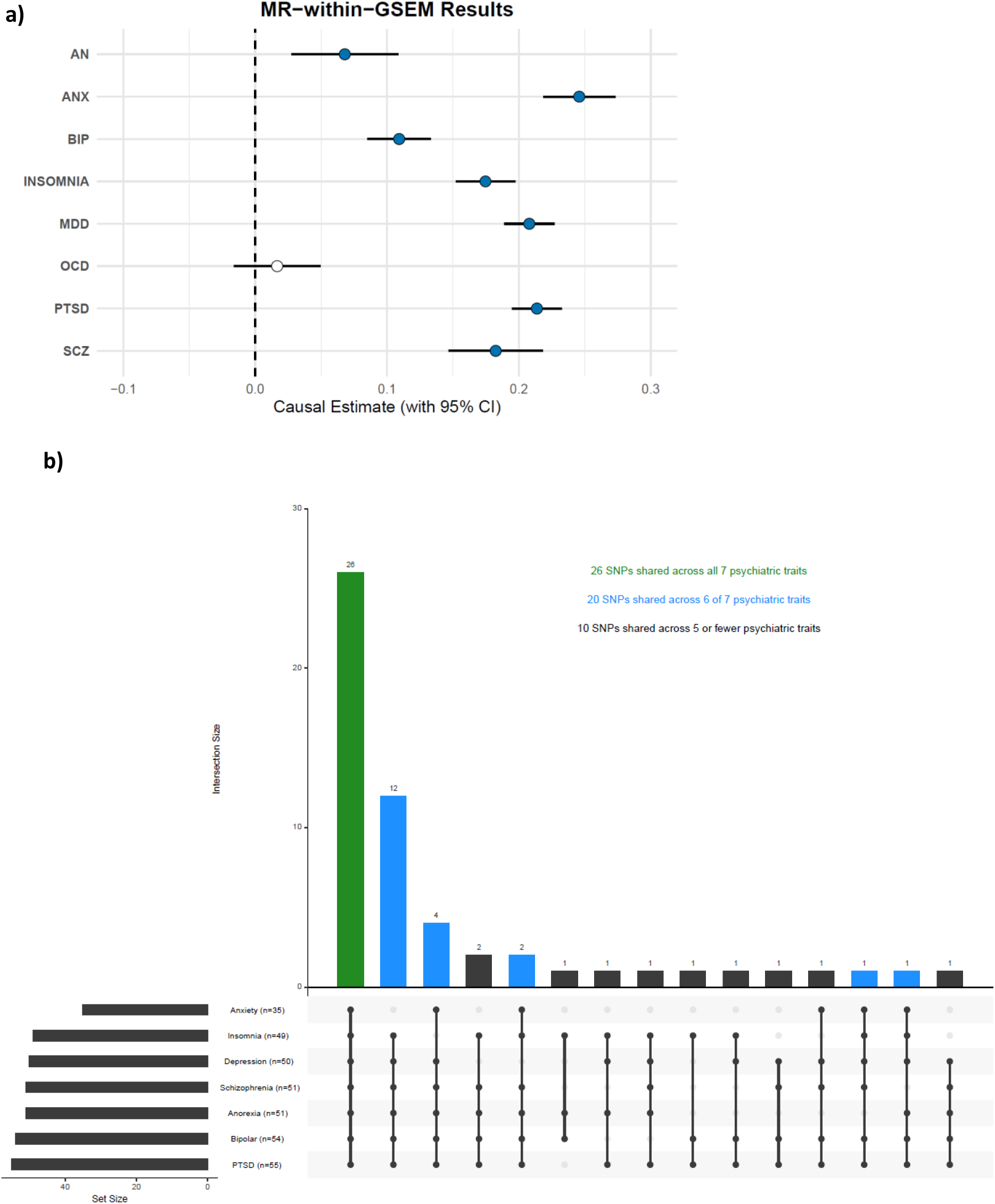
**a)** Mendelian Randomization results in genomic SEM framework. *Unique Smoking* was used as an exposure and eight psychiatric traits were used as outcomes. Filled dots represent significance at *p<*.05 after Bonferroni correction. Causal estimate is expressed as the unstandardized genomic SEM estimate. **b)** Intersection of all 56 SNPs that contributed to the relationship between *Unique Smoking* and any of the seven common psychiatric traits causally associated with *Unique Smoking*.

## Discussion

This study examined the relationship between tobacco smoking and psychiatric outcomes by combining summary-level data of genome-wide association studies in multivariate models in Genomic SEM. By jointly modelling tobacco smoking, substance use, and socioeconomic status (SES)-related traits, we first aimed to isolate genetic liability that is more specific to smoking behaviour and less confounded by broader substance use or socioeconomic pathways. We found that tobacco smoking contains a substantial genetic component that is independent from the shared genetic liability of other substance use and SES-related traits. Next, after accounting for the shared genetic architecture with the other variables, we found strong evidence for a causal effect of smoking on several psychiatric outcomes, confirming previous studies.

The hierarchical Genomic SEM model showed that smoking-related traits share considerable genetic overlap with both other types of substance use and socioeconomic factors, but also retain a distinct smoking-specific component. These results align with previous studies that used similar approaches to tease apart shared genetic architecture from unique components (40–43). These results also align with literature showing that genetic studies of tobacco smoking are partially confounded by SES and broader substance use liability (20, 21, 44). We further extended this work by characterizing the genetic variants that are unique to tobacco smoking. We found 121 independent genome-wide significant hits, several of them in nicotine receptor and risk-taking genes that were previously reported. Gene-based and gene-set enrichment analyses additionally implicated genes regulating brain morphology, connectivity, and structure, supporting the theory that brain changes may be involved in the association between smoking and mental illness (45–48). In addition, genetic correlations modelled in Genomic SEM showed that the *Unique Smoking Factor* resembled nicotine dependence and heaviness of use more than it did initiation, cessation, or use disorder. This pattern is consistent with the *Unique Smoking Factor* capturing genetic liability that is more biologically proximal to the pharmacological effects of nicotine, whereas initiation and cessation are more strongly shaped by social and environmental factors, including the substance use and SES confounders removed from this factor. As expected, the substance use and SES related traits were less correlated with the non-overlapping, *Unique Smoking Factor* than with the overlapping general one. Together with the fit statistics, these results show that we were able to derive a refined smoking latent factor. We then performed a GWAS on this factor and used the derived summary level data for smoking to examine the genetic and causal associations with psychiatric outcomes.

Attenuation of genetic correlations between the *Unique Smoking Factor* and psychiatric outcomes when compared to the *GSMK Factor,* provided evidence that the correlation between smoking and mental health partially reflects liability pathways shared with SES and substance use factors rather than smoking-specific biology alone. Such pathways include important confounding factors, such as socioeconomic status and broader substance use liability in this project, but may also include impulsivity-related traits, or other correlated behavioural dimensions. This interpretation is broadly consistent with previous literature indicating that the correlation between smoking and psychiatric disorders is partially mediated by broader externalizing, socioeconomic, or behavioural domains (30, 49–52). Nevertheless, MDD, PTSD, and bipolar disorder retained a significant genetic correlation with the *Unique Smoking Factor*. This persistence suggests that they may share more direct biological pathways with tobacco smoking than other psychiatric outcomes, although genetic correlation alone cannot distinguish whether this reflects shared underlying biology or a genuine causal effect of smoking on these outcomes. Several mechanisms have been reported as potential drivers of the overlap between smoking and psychiatric outcomes, including nicotine-related modulation of dopaminergic and serotonergic systems, inflammatory pathways, or shared neurobiological vulnerability to stress responsivity and reward dysregulation (53–55).

Despite the attenuation of genetic correlations when accounting for overlapping variance with substance use and SES factors, Mendelian Randomization analyses continued to support a causal effect of smoking on most psychiatric outcomes examined. Significant effects were observed for all disorders, with the strongest estimates found for anxiety disorders, MDD, and PTSD. Notably, no significant causal effect was observed for OCD, a novel finding given that this exposure-outcome relationship has not, to our knowledge, been previously examined using Mendelian randomization. These findings are broadly consistent with previous two-sample MR studies reporting detrimental causal effects of smoking on mental health outcomes (9, 31, 47, 56). The persistence of causal effects after accounting for shared liabilities and pleiotropic effects strengthens the argument that smoking itself contributes to psychiatric risk, rather than merely acting as a proxy for correlated behavioural or socioeconomic factors. Moreover, many of the variants contributing to these causal effects were shared across multiple psychiatric disorders, suggesting that smoking may influence general biological pathways underlying broad psychiatric vulnerability rather than disorder-specific mechanisms. Psychiatric disorders share substantial genetic architecture at the disorder level (57, 58), though broader confirmation of shared variant-level mechanisms across psychiatric outcomes is needed. Future research should investigate the biological processes underlying these potential shared effects and determine whether common pathways mediate the impact of smoking across psychiatric outcomes.

Several psychiatric outcomes showed non-significant genetic correlations with the *Unique Smoking Factor* while still demonstrating positive causal effects in MR analyses. These findings reflect fundamental differences between genetic correlation and causal inference. Genetic correlation summarizes shared signal as a single genome-wide average — in our framework, after conditioning on substance use and SES-related traits — and reflects what is captured in the underlying GWAS effects, including potential non-genetic sources of covariance. In contrast, MR relies on a small set of genome-wide significant instruments and tests whether genetic variants that increase smoking liability have causal effects on an outcome. Because different genomic regions can show heterogeneous, even opposing, directions of overlap that are masked in a global average (59), a psychiatric trait may show limited residual genetic overlap with smoking while still being causally affected by smoking behaviour. These findings highlight that genetic correlation and causal inference provide complementary, but distinct, sources of evidence. Importantly, the non-significant genetic correlations should not be interpreted as evidence of absence of shared genetic influence. In addition, point estimates were generally positive but imprecise, with wide confidence intervals that included zero, suggesting limited power to detect modest residual genome-wide covariance after conditioning. This is particularly relevant in our framework, where conditioning on substance use and socioeconomic traits likely reduces the magnitude of detectable shared signal.

This study highlights the utility of multivariate genetic methods for improving MR instrument specificity in psychiatric and behavioural genetics. Many complex behaviours are genetically intertwined with correlated traits, making it difficult to distinguish trait-specific biology from broader shared liability. Genomic SEM provides a framework to model this complexity explicitly and derive more refined latent constructs for downstream causal analyses. Although statistical methods such as multivariable Mendelian Randomization (MVMR) offer the possibility to adjust for multiple exposures simultaneously, their application is often limited in practice by weak or partially overlapping genetic instruments across traits, which can reduce power and lead to unstable estimates. In contrast, refinement of GWASs using Genomic SEM allows the construction of latent, conditional genetic factors that capture more specific sources of variation while leveraging genome-wide signal, potentially improving instrument specificity and interpretability in downstream causal inference studies. Realizing this benefit, however, requires that causal inference be conducted within the genomic SEM framework itself, where the model can appropriately account for the factor structure, indicator-specific residuals, and instrument screening jointly. Used this way, genomic SEM offers a principled route to improving instrument specificity, rather than a shortcut around standard MR assumptions.

Several limitations should be considered when interpreting these findings. First, the Genomic SEM model was theory-driven, not data-driven. Although extensive validation analyses supported the plausibility of the model structure, latent factors in Genomic SEM remain conditional on the specifications imposed by the researcher. Alternative model structures may partition shared and unique variance differently. Nonetheless, the model was preregistered before analyses began. This approach also has important advantages. Theory-driven specification can enhance interpretability by aligning latent constructs with prior evidence on the genetic architecture of related traits, thereby facilitating comparison across studies. It also reduces researcher degrees of freedom relative to fully data-driven approaches, which may improve transparency and replicability.

Second, although we explicitly modelled overlap with two of the most important confounders (substance use and SES-related traits), residual confounding likely remains. Smoking behaviour is highly polygenic and influenced by numerous behavioural, environmental, and social factors, including impulsivity, risk-taking, and personality traits that were not directly included in the model. Some of these factors may represent confounders, while others may constitute intrinsic components of smoking liability itself. Third, conventional MR sensitivity analyses such as MR-Egger cannot be directly implemented within the Genomic SEM framework. Although we addressed pleiotropy by modelling direct SNP effects on outcomes, this approach does not fully eliminate the possibility of residual horizontal pleiotropy. Fourth, all analyses were restricted to European ancestry GWASs due to the lack of available summary statistics of well-powered GWAS in non-European ancestries and due to methodological constraints related to matching linkage disequilibrium reference panels. Sociocultural, political, and environmental context has an impact on smoking behavior and substance use, thus it is important for future work to address this knowledge gap. Finally, reverse causality was outside the scope of this study. Extensive literature suggests that the relationship between smoking and psychiatric outcomes is likely bidirectional, with mental health problems also increasing the likelihood of smoking initiation and persistence. Our findings therefore should not be interpreted as excluding reverse pathways. While bidirectional Mendelian randomization would in principle be informative, it is less straightforward in the present framework because the Genomic SEM-derived smoking factor represents a model-based latent construct rather than an observed phenotype, and reversing the direction would require a separate and potentially non-equivalent latent specification for psychiatric liability. In addition, differences in instrument strength and the conditional nature of the latent factor complicate interpretation of reverse causal estimates.

By integrating multiple smoking phenotypes and explicitly modelling overlap with substance use and socioeconomic status, this study derived a refined genetic representation of tobacco smoking that appears more specific to smoking-related biology. Using this refined instrument, we found evidence consistent with a causal effect of smoking on multiple psychiatric outcomes even after accounting for major shared liabilities. These findings contribute both methodologically—by improving the specificity of genetic instruments for behavioural traits—and substantively, by strengthening evidence that tobacco smoking itself may play a direct role in psychiatric disease risk.

## Methods

The analysis plan for this study was preregistered on OSF (https://osf.io/xryju). Changes to the preregistration can be found in Supplementary Table 3. We conducted four sets of analyses: (1) *Latent factor architecture mapping,* to capture genetic signal for smoking overlapping with and separate from socioeconomic and substance use traits, (2) *Evaluation of the latent Unique Smoking Factor*, to ensure that the latent smoking genetic signal actually reflects smoking behaviours, (3) *Genetic correlations between the latent smoking factor and a range of mental health outcomes*, to compare the associations of the smoking-specific genetic signal with those of the signal shared with socioeconomic and substance use traits, and (4) *Examining causal associations of Unique Smoking with mental health through MR*, to test causal effects of smoking.

### Data

For this study we used summary-level data of the largest available European ancestry GWASs for each trait (Supplementary Table 1) (published before August 2025). When applicable, we requested permission from 23andMe, Inc., to use summary statistics of GWAS meta-analyses including this cohort. We restricted analyses to European-ancestry GWASs because Genomic SEM requires genetic covariance matrices to be estimated using an LD reference panel matched to a single ancestry; differing LD structures across populations preclude combining ancestries without such alignment. We screened all included GWASs for data on other ancestries, but replication of the model was not feasible due to a lack of sufficiently powered non-European GWASs for these traits (several traits with below 3% SNP-h2 and below 10 LDSC Z-scores).

We included traits with a significant SNP-based heritability larger than 3%, estimated using Linkage-Disequilibrium Score Regression (LDSC) (60). Most data were obtained from published original GWAS papers. Summary statistics for Townsend Deprivation Index (a measure of area-level socioeconomic deprivation – UKB code 189) and caffeine use (UKB code 1498) were obtained from the Westlake Yang Lab website, which conducted GWASs in the UK Biobank cohort (61) using fastGWA (62, 63). The caffeine use GWAS results were further meta-analysed with those reported by Cornelis et al. (2014) (64).

We aimed to include a broad range of traits that could be classified into one of three domains: tobacco smoking, other substance use, or socioeconomic status. In total, this resulted in the inclusion of seven tobacco-related traits, eight other substance use–related traits, and four socioeconomic status–related traits. GWAS summary statistics for eight common psychiatric disorders were additionally used for the genetic correlations analyses (step 3) and Mendelian randomization analyses (step 4). All summary statistics were pre-processed using the tidyGWAS R package (65) to scan for missing values, duplications, indels, and to match the RSID of the summary statistics with a dbSNP reference file. Informed consent was obtained originally by the participating studies from each GWAS.

### Analyses

#### 1. Latent factor architecture mapping

We used LDSC within the GenomicSEM (38) framework to assess genetic correlations and SNP-based heritability between all nineteen traits included in the model. These nineteen summary-level GWAS data were used as primary input for Genomic SEM. Genetic correlations were plotted as a heatmap using the corrplot package in R (66).

Next, we used a theory-driven hierarchical Genomic SEM approach, as previously described by Pasman et al. (30) and Grotzinger et al. (67), to capture genetic signal for smoking overlapping with and separate from socioeconomic and substance use traits. Our model consists of two layers of latent factors in a variance-covariance decomposition (See Figure 1a). The first (hierarchical) layer groups all tobacco smoking related signals into one general smoking factor (from now on ‘GSMK’), all shared substance use related genetic signal into a general substance use latent factor (from now on ‘GSU’), and all SES-related genetic signal into a SES latent factor (from now on ‘GSES’). These three factors capture genetic variation within each domain, but they still overlap genetically between domains. That overlap is what the second layer is designed to unpack. The second (hierarchical) layer decomposes these factors into two shared and one unique factor. One factor captures genetic signal shared by all first layer general latent factors (from now on referred to as ‘common SMK/SU/SES’), and one factor captures the signal shared between the general substance use (GSU) and tobacco factors (GSMK), not shared by GSES (from now on ‘common SU’). This decomposition results in a third unique factor which exclusively captures the genetic signal of tobacco smoking (from now on ‘*Unique Smoking’*) after removing all overlap with the common SMK/SU/SES and common SU factors.

Model fit was evaluated using the Comparative Fit Index (CFI) and Standardized Root Mean Square Residual (SRMR), with commonly used thresholds of CFI >0.90 and SRMR <0.10 taken as reference. However, as our model was theory-driven rather than purely data-driven — aimed at isolating the unique genetic signal of tobacco smoking from potential confounders — fit indices were interpreted with some leniency relative to these traditional thresholds, prioritizing theoretical relevance and study objectives over optimal statistical fit.

#### 2. Evaluation of the latent Unique Smoking Factor

In order to evaluate the construct validity of the *Unique Smoking Factor*, two tests were performed. First, we studied how different the *Unique Smoking Factor* was compared to GSMK. In a first step, within the Genomic SEM model, we fit genetic correlations between GSMK and the observed traits that compose it (see Figure 3), and the genetic correlations between the *Unique Smoking Factor* and these same traits and then compared them. If the *Unique Smoking Factor* genuinely isolates smoking-specific signal, it should show genetic correlations with these smoking-related traits that are comparable in magnitude to those of GSMK; a marked weakening of these correlations would suggest that the Unique Smoking Factor no longer adequately captures smoking behaviour itself. In a second step, to ensure that overlapping signal with confounders is less present in the *Unique Smoking Factor* than in the GSMK, genetic correlations between both factors (GSMK and the *Unique Smoking Factor),* and all the SES and substance use-related traits included in the model were calculated; we expected these correlations to be attenuated for the *Unique Smoking Factor* relative to GSMK, confirming that shared signal with these confounders had been successfully removed. All correlations were tested against a nominal significance threshold of α = 0.05. For comparisons between correlations (e.g., GSMK vs. Unique Smoking with the same trait), statistical significance was assessed using the delta method, in which the Jacobian of each genetic correlation with respect to the elements of the genetic covariance matrix was numerically computed and combined with the sampling covariance matrix from LDSC to derive the variance of, and a Z-test for the difference between the two correlations. Multiple testing was assessed by calculating False Discovery rate (FDR), both for the individual correlations and the comparisons (68). All results were plotted using the ggplot2 package in R.

Second, multivariate GWAS was used to identify loci specific to tobacco smoking via the Unique Smoking factor. Genomic SEM allows to estimate the effects of millions of SNPs on any identified latent factor, virtually running a GWAS on the latent factor. We used the userGWAS function of Genomic SEM to run a GWAS on the *Unique Smoking Factor*. To further characterize the biological relevance of the loci associated with the *Unique Smoking Factor*, Functional Mapping and Annotation of Genome-Wide Association Studies (FUMA) (69) was applied. FUMA provides a structured biological interpretation by integrating positional, functional, and expression-based information. First, we identified independent risk loci (at *R*^2^ <0.01 and distance >250 kb). After listing the top SNP associations, we conducted gene-based association tests in MAGMA (70) using FUMA. Gene-based tests aggregate SNP-level effects into gene-level signals, allowing us to identify genes whose combined variants show significant association with the *Unique Smoking Factor*. We also carried out gene-set enrichment analyses to evaluate whether these genes cluster in known biological pathways or functional categories, providing clues about the underlying molecular mechanisms. Finally, the same framework was used to test whether gene-level effects correlate with gene expression levels in 53 GTEx tissue types, including various brain regions, metabolic tissues, cardiovascular tissues, and others. This step helps determine whether the genetic architecture of the *Unique Smoking Factor* aligns with biological activity in specific tissues or systems.

#### 3. Genetic correlations between the latent smoking factor and a range of mental health outcomes

To better understand the role of smoking in mental health outcomes, we first examined the genetic correlations between the *Unique Smoking Factor* and eight common psychiatric disorders, and compared them to the correlations of these traits with GSMK. The mental health outcomes were selected based on their global prevalence and prior evidence of associations with smoking in the literature. Insomnia was included as an outcome due to its high prevalence and its transdiagnostic relevance across psychiatric disorders. Summary statistics for seven psychiatric disorders were selected from the Psychiatric Genetics Consortium (PGC) - prioritising the largest, most recent GWAS, while the summary statistics of insomnia came from Watanabe et al. (71). Details are presented in Supplementary Table 1.

Genetic correlations were calculated within Genomic SEM by modelling each psychiatric trait as a latent factor and allowing this factor to correlate to the *Unique Smoking Factor* in one iteration and to GSMK in another (See Figure 1b). The mental health traits are first loaded to latent factors to avoid scaling problems derived from calculating correlations between indicators in different hierarchical layers.

#### 4. Examining causal associations of the Unique Smoking Factor with mental health through MR

Lastly, we assessed the potential causal effect of smoking on mental health outcomes using an MR approach implemented within the Genomic SEM framework. Unlike conventional two-sample MR, which estimates causal effects using SNP-exposure and SNP-outcome associations from separate GWASs, this approach models genetic effects on exposure and outcomes jointly at the genome-wide level, while explicitly accounting for sample overlap, differences in sample size, heritability, and residual population stratification (38, 67).

MR is a statistical method that uses genetic variants as instrumental variables to infer causal effects of an exposure on an outcome and relies on three core assumptions; genetic variants being used as an instrument: (1) must be reliably associated with the exposure of interest (relevance), (2) must be independent from confounders of the exposure-outcome association (independence), and (3) must not influence the outcome in any other way than through the exposure (exclusion restriction). Given the complexity of smoking behaviour and psychiatric traits, violations of these assumptions are plausible, making careful instrument selection and sensitivity analyses essential.

In this study, the potential causal relationship between smoking and psychiatric traits was modelled unidirectionally since our interest lies in the effect of smoking on mental health outcomes. Genome-wide significant (p < 5 × 10⁻⁸), independent (r² < 0.01) SNPs associated with the *Unique Smoking Factor* - obtained from the Genomic SEM GWAS – were selected as genetic instruments. These SNPs were clumped and harmonized with each psychiatric outcome using the TwoSampleMR R package (72). This process resulted in eight sets of harmonized SNPs, one for each pair of exposure-psychiatric outcome.

Causal effects were then estimated by modelling the SNPs through the *Unique Smoking Factor* onto each psychiatric trait within Genomic SEM, following the procedures described by Lukas et al. (73) and Grotzinger et al. (67). Genomic SEM accounts for sample overlap, differences in sample size and heritability across GWASs, and partially accounts for residual population stratification present in the summary statistics. In addition, this framework allows for confounding genetic influences - in our model with shared genetic variance with other substance use traits or socioeconomic status - to be explicitly modelled, thereby partially addressing the independence assumption.

Since MR is embedded within the Genomic SEM framework, conventional MR sensitivity analyses for horizontal pleiotropy (e.g. MR-Egger) are not directly applicable. Instead, pleiotropy was addressed by iteratively modelling direct SNP loadings on the psychiatric outcomes. SNPs that no longer showed a significant effect on the outcome after accounting for these direct pathways were excluded. SNPs retaining nominal significance (p < 0.05) after accounting for direct pathways were retained as instruments consistent with the hypothesized causal effect of smoking on the outcome. Mean F statistic, mean R^2^, and total R^2^ of the exposure were calculated.

Because these analyses tested the causal effect of smoking on 8 psychiatric outcomes, a Bonferroni correction was applied across these 8 tests to guard against false positives, with α = 0.05/8 = 0.00625 taken as the corrected significance threshold for each outcome-level MR result.

Finally, to examine whether individual SNPs exerted effects across multiple psychiatric outcomes or were outcome-specific, instrument SNPs were compared across traits and their overlap visualized using the UpSetR package in R.

## Data availability

GWAS summary data is available in the original study papers. Permission was obtained from 23andMe Research Institute and dbGaP when needed. The dbGaP individual-level data are accessible to all researchers upon application. The full GWAS summary statistics for the 23andMe discovery data set will be made available through 23andMe to qualified researchers under an agreement with 23andMe that protects the privacy of the consented 23andMe participants. Please visit https://research.23andme.com/collaborate/#dataset-access/ for more information and to apply to access the data.

## Code availability

All code and scripts used for this project are shared on our GitHub, GitHub-AdriaTunezAquilue/Smoking_Genomic_SEM.

## Data Availability

GWAS summary data is available in the original study papers. Permission was obtained from 23andMe Research Institute and dbGaP when needed. The dbGaP individual-level data are accessible to all researchers upon application. The full GWAS summary statistics for the 23andMe discovery data set will be made available through 23andMe to qualified researchers under an agreement with 23andMe that protects the privacy of the consented 23andMe participants.

## Acknowledgements

AT, APSO, and KJHV are supported by the Dutch Research Council (NWO), Open Competition grant (project number 406.22.GO.043). KJHV is supported by the Foundation Volksbond Rotterdam.

JAP is supported by a 2024 BBRF Distinguished Investigator Grant (32124) from the Brain & Behavior Research Foundation.

AA is supported by the Amsterdam UMC Fellowship.

DJAS is supported by NWO open competition L project 406.23.PPO.050.

JLT is funded by the European Union (ERC, UNRAVEL-CAUSALITY, 101076686). Views and opinions expressed are however those of the author(s) only and do not necessarily reflect those of the European Union or the European Research Council. Neither the European Union nor the granting authority can be held responsible for them.

We would like to thank the research participants and employees of 23andMe, Inc., for making this work possible. We also thank the research participants and employees of the UK Biobank, for which we have access under project application 509769. We gratefully acknowledge the Psychiatric Genomics Consortium contributing studies and thank the participants in those studies without whom this effort would not have been possible. We thank SURFsara for the usage of the Snellius cluster computer.

## Author contributions

KJHV, JAP, DJAS, JLT, and AT were involved in the conception and design of the study. AT was involved in data analysis. All authors were involved in writing the manuscript.

## References

1. Peacock A, Leung J, Larney S, Colledge S, Hickman M, Rehm J, et al. Global statistics on alcohol, tobacco and illicit drug use: 2017 status report. Addiction. 2018;113(10).

2. Physicians RCo. Smoking and mental health. 2013.

3. National Survey of Drug Use and Health (NSDUH) Releases. CBHSQ Data; 2019.

4. Organization WH. The vicious cycle of tobacco use and mental illness – a double burden on health. World Health Organization. 2021.

5. Leon Jd, Diaz FJ. A meta-analysis of worldwide studies demonstrates an association between schizophrenia and tobacco smoking behaviors. Schizophrenia Research. 2005;76(2-3).

6. Sagud M, Mihaljevic Peles A, Pivac N. Smoking in schizophrenia: recent findings about an old problem. Current Opinion in Psychiatry. 2019;32(5).

7. Khantzian EJ. The Self-Medication Hypothesis of Substance Use Disorders: A Reconsideration and Recent Applications. Harvard Review of Psychiatry. 1997;4(5).

8. Taylor G, McNeill A, Girling A, Farley A, Lindson-Hawley N, Aveyard P. Change in mental health after smoking cessation: systematic review and meta-analysis. BMJ. 2014;348(1).

9. Taylor GMJ, Treur JL. An application of the stress-diathesis model: A review about the association between smoking tobacco, smoking cessation, and mental health. International Journal of Clinical and Health Psychology. 2023;23(1).

10. Treur JL, Munafò MR, Logtenberg E, Wiers RW, Verweij KJH. Using Mendelian randomization analysis to better understand the relationship between mental health and substance use: a systematic review. Psychological Medicine. 2021;51(10).

11. Kleinman RA, Barnett BS. Smoking Cessation as a Priority for Psychiatrists. JAMA Psychiatry. 2024;81(10).

12. Teasdale SB, Machaczek KK, Marx W, Eaton M, Chapman J, Milton A, et al. Implementing lifestyle interventions in mental health care: third report of the Lancet Psychiatry Physical Health Commission. The Lancet Psychiatry. 2025;12(9).

13. Plurphanswat N, Kaestner R, Rodu B. The Effect of Smoking on Mental Health. American Journal of Health Behavior. 2017;41(4).

14. Abdellaoui A, Yengo L, Verweij KJH, Visscher PM. 15 years of GWAS discovery: Realizing the promise. The American Journal of Human Genetics. 2023;110(2).

15. George Davey Smith, Shah E. ‘Mendelian randomization’: can genetic epidemiology contribute to understanding environmental determinants of disease? International Journal of Epidemiology. 2003;32(1).

16. Larsson SC, Burgess S. Appraising the causal role of smoking in multiple diseases: A systematic review and meta-analysis of Mendelian randomization studies. eBioMedicine. 2022;82.

17. Firth J, Solmi M, Wootton RE, Vancampfort D, Schuch FB, Hoare E, et al. A meta-review of “lifestyle psychiatry”: the role of exercise, smoking, diet and sleep in the prevention and treatment of mental disorders. World Psychiatry. 2020;19(3).

18. Abdellaoui A, Verweij KJH. Dissecting polygenic signals from genome-wide association studies on human behaviour. Nature Human Behaviour. 2021;5(6).

19. Pasman JA, Verweij KJH, Gerring Z, Stringer S, Sanchez-Roige S, Treur JL, et al. GWAS of lifetime cannabis use reveals new risk loci, genetic overlap with psychiatric traits, and a causal effect of schizophrenia liability. Nature Neuroscience. 2018;21(9).

20. Marees AT, Smit DJA, Abdellaoui A, Nivard MG, van den Brink W, Denys D, et al. Genetic correlates of socio-economic status influence the pattern of shared heritability across mental health traits. Nature Human Behaviour. 2021;5(8).

21. Verweij KJH, Treur JL, Vink JM. Investigating causal associations between use of nicotine, alcohol, caffeine and cannabis: a two-sample bidirectional Mendelian randomization study. Addiction. 2018;113(7).

22. Watanabe K, Stringer S, Frei O, Umićević Mirkov M, de Leeuw C, Polderman TJC, et al. A global overview of pleiotropy and genetic architecture in complex traits. Nat Genet. 2019;51(9).

23. Nivard MG, Verweij KJH, Minică CC, Treur JL, Vink JM, Boomsma DI. Connecting the dots, genome-wide association studies in substance use. Mol Psychiatry. 2016;21(6).

24. Abdellaoui A, Dolan CV, Verweij KJH, Nivard MG, Abdellaoui A, Dolan CV, et al. Gene– environment correlations across geographic regions affect genome-wide association studies. Nat Genet. 2022;54(9).

25. Cambron C, Kosterman R, Hawkins JD. Neighborhood Poverty Increases Risk for Cigarette Smoking From Age 30 to 39. Annals of Behavioral Medicine. 2019;53(9).

26. Karriker-Jaffe KJ. Neighborhood socioeconomic status and substance use by U.S. adults. Drug and Alcohol Dependence. 2013;133(1).

27. Reed ZE, Wootton RE, Khouja JN, Richardson TG, Sanderson E, Smith GD, et al. Exploring pleiotropy in Mendelian randomisation analyses: What are genetic variants associated with ‘cigarette smoking initiation’ really capturing? Genetic Epidemiology. 2025;49(1).

28. Burgess S, Davey Smith G. How humans can contribute to Mendelian randomization analyses. International Journal of Epidemiology. 2019;48(3).

29. Song W, Lin GN, Yu S, Zhao M. Genome-wide identification of the shared genetic basis of cannabis and cigarette smoking and schizophrenia implicates NCAM1 and neuronal abnormality. Psychiatry Research. 2022;310.

30. Pasman JA, Allegrini AG, Tunez A, Abdellaoui A, Smit DJA, Nivard MG, et al. A Systematic Investigation of the Common Genetic Architecture of Substance Use Traits and the Relationship with Mental Health. European Addiction Research. 2025.

31. Wootton RE, Richmond RC, Stuijfzand BG, Lawn RB, Sallis HM, Taylor GMJ, et al. Evidence for causal effects of lifetime smoking on risk for depression and schizophrenia: a Mendelian randomisation study. Psychological Medicine. 2020;50(14).

32. Saunders GRB, Wang X, Chen F, Jang S-K, Liu M, Wang C, et al. Genetic diversity fuels gene discovery for tobacco and alcohol use. Nature. 2022;612(7941).

33. Toikumo S, Jennings MV, Pham BK, Lee H, Mallard TT, Bianchi SB, et al. Multi-ancestry meta-analysis of tobacco use disorder identifies 461 potential risk genes and reveals associations with multiple health outcomes. Nature Human Behaviour. 2024;8(6).

34. Johnson EC, Lai D, Balbona JV, Miller AP, Hatoum AS, Deak JD, et al. Multi-ancestral genome-wide association study of clinically defined nicotine dependence reveals strong genetic correlations with other substance use disorders and health-related traits. Psychological Medicine. 2025;55.

35. Sullivan PF, Daly MJ, O’Donovan M, Sullivan PF, Daly MJ, O’Donovan M. Genetic architectures of psychiatric disorders: the emerging picture and its implications. Nature Reviews Genetics. 2012;13(8).

36. Vink JM, Smit AB, Geus EJCd, Sullivan P, Willemsen G, Hottenga J-J, et al. Genome-wide Association Study of Smoking Initiation and Current Smoking. The American Journal of Human Genetics. 2009;84(3).

37. Liu M, Jiang Y, Wedow R, Li Y, Brazel DM, Chen F, et al. Association studies of up to 1.2 million individuals yield new insights into the genetic etiology of tobacco and alcohol use. Nat Genet. 2019;51(2).

38. Grotzinger AD, Rhemtulla M, de Vlaming R, Ritchie SJ, Mallard TT, Hill WD, et al. Genomic structural equation modelling provides insights into the multivariate genetic architecture of complex traits. Nature Human Behaviour. 2019;3(5).

39. Lee PH, Anttila V, Won H, Feng Y-CA, Rosenthal J, Zhu Z, et al. Genomic Relationships, Novel Loci, and Pleiotropic Mechanisms across Eight Psychiatric Disorders. Cell. 2019;179(7).

40. Cao Y, Yang X, Svensson P, Chung Wen RW, Han Sng TJ, Islam I, et al. Genetic subtraction reveals divergent pathways and targets in anxiety-related and anxiety-independent TMD. The Journal of Headache and Pain. 2026;27(1).

41. Demange PA, Malanchini M, Mallard TT, Biroli P, Cox SR, Grotzinger AD, et al. Investigating the genetic architecture of noncognitive skills using GWAS-by-subtraction. Nat Genet. 2021;53(1).

42. Pasman JA, Demange PA, Guloksuz S, Willemsen AHM, Abdellaoui A, ten Have M, et al. Genetic Risk for Smoking: Disentangling Interplay Between Genes and Socioeconomic Status. Behavior Genetics. 2021;52(2).

43. Vries LPd, Demange PA, Baselmans BML, Vinkers CH, Pelt DHM, Bartels M. Distinguishing happiness and meaning in life from depressive symptoms: A GWAS-by-subtraction study in the UK Biobank. American Journal of Medical Genetics. 2024;195(1).

44. Albayrak M, Turhan K. Genetic and Environmental Factors of Nicotine Addiction: Examination of Multiple Substance Addictions. American Journal of Medical Genetics. 2026;201(2).

45. Boer OD, El Marroun H, Muetzel RL, Boer OD, El Marroun H, Muetzel RL. Adolescent substance use initiation and long-term neurobiological outcomes: insights, challenges and opportunities. Molecular Psychiatry 2024 29:7. 2024;29(7).

46. Bruijnzeel AW. Tobacco addiction and the dysregulation of brain stress systems. Neuroscience & Biobehavioral Reviews. 2012;36(5).

47. Chen Y, Ma X, Yin Y, Wu Y, Huang Y, Tang Y, et al. Causal Associations Between Smoking, Brain Structural Alterations and Psychiatric Disorders: Evidence From a Mediation Analysis. Addiction Biology. 2025;30(12).

48. Lin F, Wu G, Zhu L, Lei H. Heavy smokers show abnormal microstructural integrity in the anterior corpus callosum: A diffusion tensor imaging study with tract-based spatial statistics. Drug and Alcohol Dependence. 2013;129(1-2).

49. Marti CN, Arora S, Loukas A. Depressive symptoms predict trajectories of electronic delivery nicotine systems, cigarette, and cannabis use across 4.5 years among college students. Addictive Behaviors. 2023;146.

50. Halladay J, Kershaw S, Devine EK, Grummitt L, Visontay R, Lynch SJ, et al. Covariates in studies examining longitudinal relationships between substance use and mental health problems among youth: A meta-epidemiologic review. Drug and Alcohol Dependence. 2025;271.

51. Chung JYC, Lim CCW, Connor JP, Hall W, Stjepanović D, Chan GCK. Examining the Relationship of Cannabis use Patterns, Mental Health, and Sociodemographic Factors: A Focus on Cannabis Vaping, Smoking and Dual-Use. Addictive Behaviors. 2025;163.

52. Berg N, Piirtola M, Marttunen M, Latvala A, Kiviruusu O. Trajectories of Concurrent Psychological Distress, Heavy Episodic Drinking and Daily Cigarette Smoking From Adolescence to Midlife: Patterns and Their Sociodemographic Correlates. Nordisk Alkohol-& Narkotikatidskrift : NAT. 2026;43(1).

53. Proud EK, Rodríguez-Ruiz M, Gummerson DM, Vanin S, Hardy DB, Rushlow WJ, et al. Chronic nicotine exposure induces molecular and transcriptomic endophenotypes associated with mood and anxiety disorders in a cerebral organoid neurodevelopmental model. Frontiers in Pharmacology. 2024;15.

54. Yan B, Hu J, Xi J, Luo L, Zheng X, Luo S, et al. The role of C-reactive proteins in tobacco smoke exposure and the risk of depression. Psychology, Health & Medicine. 2025.

55. Zhou Y, Zhuang J, Bian Q, Xu Z, Chen Y, Yang H, et al. METS-IR and SII as mediators in the association between smoking and depressive symptoms: insights from NHANES (2005–2018). BMC Psychiatry. 2025;25(1).

56. Vermeulen JM, Wootton RE, Treur JL, Sallis HM, Jones HJ, Zammit S, et al. Smoking and the risk for bipolar disorder: evidence from a bidirectional Mendelian randomisation study. The British Journal of Psychiatry. 2021;218(2).

57. Williams CM, Peyre H, Wolfram T, Lee YH, Seidlitz J, Ge T, et al. Characterizing the phenotypic and genetic structure of psychopathology in UK Biobank. Nature Mental Health. 2024;2(8).

58. Grotzinger AD, Werme J, Peyrot WJ, Frei O, de Leeuw C, Bicks LK, et al. Mapping the genetic landscape across 14 psychiatric disorders. Nature. 2025;649(8096).

59. Werme J, van der Sluis S, Posthuma D, de Leeuw CA, Werme J, van der Sluis S, et al. An integrated framework for local genetic correlation analysis. Nat Genet. 2022;54(3).

60. Bulik-Sullivan BK, Loh P-R, Finucane HK, Ripke S, Yang J, Patterson N, et al. LD Score regression distinguishes confounding from polygenicity in genome-wide association studies. Nat Genet. 2015;47(3).

61. Bycroft C, Freeman C, Petkova D, Band G, Elliott LT, Sharp K, et al. The UK Biobank resource with deep phenotyping and genomic data. Nature. 2018;562(7726).

62. Jiang L, Zheng Z, Qi T, Kemper KE, Wray NR, Visscher PM, et al. A resource-efficient tool for mixed model association analysis of large-scale data. Nat Genet. 2019;51(12).

63. Abdellaoui A, Sanchez-Roige S, Sealock J, Treur JL, Dennis J, Fontanillas P, et al. Phenome-wide investigation of health outcomes associated with genetic predisposition to loneliness. Human Molecular Genetics. 2019;28(22).

64. Cornelis MC, Byrne EM, Esko T, Nalls MA, Ganna A, Paynter N, et al. Genome-wide meta-analysis identifies six novel loci associated with habitual coffee consumption. Mol Psychiatry. 2014;20(5).

65. Harder A. tidyGWAS: Quality control for GWAS summary statistics with minimal filtering. 2025.

66. Wei T, Simko V. R package ’corrplot’: Visualization of a Correlation Matrix. 2024.

67. Grotzinger AD, Mallard TT, Akingbuwa WA, Ip HF, Adams MJ, Lewis CM, et al. Genetic architecture of 11 major psychiatric disorders at biobehavioral, functional genomic and molecular genetic levels of analysis. Nat Genet. 2022;54(5).

68. Morgan GB, Hodge KJ, Wells KE, Watkins MW, Morgan GB, Hodge KJ, et al. Are Fit Indices Biased in Favor of Bi-Factor Models in Cognitive Ability Research?: A Comparison of Fit in Correlated Factors, Higher-Order, and Bi-Factor Models via Monte Carlo Simulations. Journal of Intelligence. 2015;3(1).

69. Watanabe K, Taskesen E, van Bochoven A, Posthuma D, Watanabe K, Taskesen E, et al. Functional mapping and annotation of genetic associations with FUMA. Nat Commun. 2017;8(1).

70. Leeuw CAd, Mooij JM, Heskes T, Posthuma D. MAGMA: Generalized Gene-Set Analysis of GWAS Data. PLOS Computational Biology. 2015;11(4).

71. Watanabe K, Jansen PR, Savage JE, Nandakumar P, Wang X, Hinds DA, et al. Genome-wide meta-analysis of insomnia prioritizes genes associated with metabolic and psychiatric pathways. Nat Genet. 2022;54(8).

72. Hemani G, Zheng J, Elsworth B, Wade KH, Haberland V, Baird D, et al. The MR-Base platform supports systematic causal inference across the human phenome. eLife. 2018;7.

73. Lukas E, Veeneman RR, Smit DJA, Ahluwalia TS, Vermeulen JM, Pathak GA, et al. A genetic exploration of the relationship between posttraumatic stress disorder and cardiovascular diseases. Transl Psychiatry. 2025;15(1).

