## Supplementary Information for "Genetic and Causal Associations Between Tobacco Smoking and Mental Health After Accounting for General Substance Use and Socioeconomic Factors"

**Content**


Supplementary Methods 2

Supplementary Figures 2

Supplementary Tables 3

References 6

### Supplementary Methods

#### Genome-wide association studies in UK Biobank

#### Townsend Deprivation Index

Because the largest published GWAS was smaller than the sample size in the UK Biobank, we downloaded the GWAS for Townsend deprivation index from the Yang Lab website. Townsend deprivation index is based on the national census output areas. Each participant is assigned a score corresponding to the output area in which their postcode is located (UK Biobank data field 189/22189). GWAS analyses were conducted in fastGWA from GCTA (Jiang et al., 2019).

#### Caffeine Use

Because the largest published GWAS was smaller than the sample size in the UK Biobank, we downloaded the GWAS for caffeine use from the Yang Lab website. Caffeine use was measured as the amount of cups of coffee drunk per day (including decaffeinated coffee) (data field 1498). Results were meta-analyzed with summary statistics from Cornelis et al. (2014) following guidelines of the METAL software (Willer et al., 2010). Results showed 5 lead ($R^2 \leq 0.1$) SNPs at chromosomes 1, 2, 8, and 15. The SNP-based heritability was $h^2_{SNP}=0.042$ (Z=11.7).

### Supplementary Figures

(Figure available in GWAScatalog_FUMA.pdf)

**Supplementary Figure 1.** Enrichment of input genes in Gene Sets, GWAS catalog reported genes. FUMA results for the *Unique Smoking Latent Factor*.

### Supplementary Tables

| **Author and Year** | **Trait** | **SNP heritability** | **Z-score** | **N effective** |
|---|---|---|---|---|
| Saunders et al. (2022) | Smoking Initiation (SmkIni) | 0.08 | 36.5 | 2630947 |
| Saunders et al. (2022) | Cigarettes per day (Cig per day) | 0.08 | 22.4 | 618489 |
| Saunders et al. (2022) | Smoking Cessation (SmkCes) | 0.05 | 29 | 894247 |
| Saunders et al. (2022) | Age at Smoking Initiation (Age SmkIni) | 0.05 | 25.5 | 618541 |
| Quach et al. (2020) | Nicotine Dependence (NicDep) | 0.09 | 7 | 46213 |
| Toikumo et al. (2024) | Tobacco Use Disorder (TUD) | 0.09 | 20.2 | 532367 |
| Johnson et al. (2025) | DSM-based Nicotine Dependence (DSM NicDep) | 0.07 | 4.2 | 47123 |
| Sanchez-Roige et al. (2018) | Alcohol use disorder symptoms (AUDIT) | 0.09 | 16.4 | 141932 |
| Saunders et al. (2022) | Alcohol; Drinks per week (Drnk per week) | 0.04 | 27.4 | 2428851 |
| Walters et al. (2018) | Alcohol Dependence (AlcDep) | 0.09 | 6.8 | 34780 |
| *Own meta-analysis** | Caffeine Use (CaffUse) | 0.03 | 9 | 540000 |
| Pasman et al. (2018) | Cannabis – Ever use (CannUse) | 0.11 | 16 | 180934 |
| Levey et al. (2023) | Cannabis Use Disorder (CUD) | 0.07 | 19 | 161053 |
| Gaddis et al. (2022) | Opioid Dependence (OpiDep) | 0.11 | 10.4 | 88114 |
| Sanchez-Roige et al. (2021) | Problematic Opioid Prescription use (ProbOpi) | 0.04 | 7.4 | 93587 |
| *Own GWAS on UKB** | Townsend Index (DeprivIdx) | 0.14 | 19 | 112151 |
| Okbay et al. (2022) | Educational Attainment (EducAtt) | 0.12 | 42.7 | 3037499 |
| Kweon et al. (2025) | Income (IncomeF) | 0.07 | 29 | 668288 |
| Akimova et al. (2024) | Occupational Status and Prestige (OccStatus) | 0.14 | 26.6 | 273157 |
| Adams et al. (2025) | Major Depressive Disorder | 0.05 | 32.4 | 1309348 |
| Trubetskoy et al. (2022) | Schizophrenia | 0.38 | 29.8 | 126282 |
| O'Connell et al. (2025) | Bipolar's Disorder | 0.13 | 27.2 | 220416 |
| Strom et al. (2026) | Anxiety | 0.07 | 23.7 | 419113 |
| Watson et al. (2019) | Anorexia Nervosa | 0.25 | 16.7 | 52042 |
| Nievergelt et al. (2024) | Post-Traumatic Stress Disorder (PTSD) | 0.07 | 27.7 | 487029 |
| Strom et al. (2025) | Obsessive-Compulsive Disorder (OCD) | 0.12 | 17.1 | 92032 |
| Watanabe et al. (2022) | Insomnia | 0.06 | 22 | 313750 |
| Hatoum et al. (2023) | Addiction | 0.02 | 22.6 | 1025550 |

**Supplementary Table 1.** GWAS summary statistics of all traits used for both genomic SEM analyses. And 8 major psychiatric traits. * Methods and results for the GWASs we conducted in UK-Biobank are reported above.

| lhs | op | rhs | Unstand_Est | Unstand_SE | STD_Genotype | STD_Genotype_SE | STD_All | p_value |
|---|---|---|---|---|---|---|---|---|
| GSES | =~ | SES | 0.261936450 | 0.00447619113170386 | 0.90183641 | 0.0154113425084015 | 0.90183641 | < 5e-300 |
| GSES | =~ | SUB | -0.100812702 | 0.00480168974632407 | -0.41413095 | 0.0197249761527894 | -0.44320235 | 7.24725056803028e-98 |
| GSES | =~ | SMK | -0.100952421 | 0.00366181139698342 | -0.35686687 | 0.0129445336040219 | -0.72408504 | 2.6207784838187e-167 |
| GSUB | =~ | SUB | 0.203903772 | 0.00728246231607011 | 0.83762175 | 0.0299157994675517 | 0.89642160 | 1.65745075263112e-172 |
| GSUB | =~ | SMK | 0.069632266 | 0.00423338486458983 | 0.24615004 | 0.0149650230484906 | 0.49943992 | 8.61606561414419e-61 |
| GSMK | =~ | SMK | 0.066318022 | 0.00374450402891985 | 0.23443429 | 0.0132368226391206 | 0.47566860 | 3.46347331023111e-70 |
| SMK | =~ | ASI | -1.096952112 | 0.0419059331268603 | -1.47514995 | 0.0563539623216543 | -0.72703101 | 4.917281428176e-151 |
| SMK | =~ | SI | 2.021647595 | 0.0792321076629584 | 1.52270544 | 0.0596777223286174 | 0.75046869 | 1.32637282337305e-143 |
| SMK | =~ | SC | 1.402497434 | 0.0512612983028934 | 1.47537049 | 0.053924882683439 | 0.72713950 | 8.2629504887024e-165 |
| SMK | =~ | NICDEP | 1.360059966 | 0.0742376064363253 | 1.31738079 | 0.0719080045206296 | 0.64927350 | 5.69308883058301e-75 |
| SMK | =~ | TUD | 1.104402533 | 0.0437191094363635 | 1.59039877 | 0.0629579132058816 | 0.78383187 | 8.50503404760377e-141 |
| SMK | =~ | DSM_NICDEP | 1.878294826 | 0.0917729548570679 | 1.85794620 | 0.0907786040601914 | 0.91569581 | 4.26039884823427e-93 |
| SUB | =~ | ALCDEP | 1.337175252 | 0.0682458383896342 | 1.06393286 | 0.0543002265961283 | 0.99415494 | 1.75666029675873e-85 |
| SUB | =~ | OPIDEP | 1.370896822 | 0.0945938911020165 | 1.06409531 | 0.0734240857229211 | 0.99429887 | 1.35240543861074e-47 |
| SUB | =~ | AUDIT | 0.197392067 | 0.0347703475763091 | 0.16349283 | 0.0287989990482216 | 0.15276888 | 1.37058956795546e-08 |
| SUB | =~ | CAN_USE | 0.332986559 | 0.0414973151918134 | 0.25405104 | 0.0316601975613786 | 0.23738696 | 1.02113457923495e-15 |
| SUB | =~ | PROB_OPI | 0.715410434 | 0.0475451717522019 | 0.95049884 | 0.0631688630613504 | 0.88814592 | 3.61451398065638e-51 |
| SUB | =~ | DPW | 0.130789785 | 0.0155922435376303 | 0.15398503 | 0.0183574692091007 | 0.14388454 | 4.93930619759047e-17 |
| SES | =~ | TOWNSEND | -0.556479770 | 0.0190036621395148 | -0.91184555 | 0.0311393347273764 | -0.82233497 | 1.71924546323862e-188 |
| SES | =~ | INCOME_F | 0.854114237 | 0.0137856803623763 | 0.91815584 | 0.0148193366659649 | 0.82802626 | < 5e-300 |
| SES | =~ | OCCUPATION | 1.253189351 | 0.0232195455689566 | 0.94897925 | 0.0175830353674631 | 0.85582366 | < 5e-300 |
| ASI | ~~ | ASI | 0.020861137 | 0.00131360035058237 | 0.47142566 | 0.0296851408186789 | 0.47142591 | 8.59558738203339e-57 |
| CPD | ~~ | CPD | 0.060585652 | 0.00301297785822189 | 0.75709740 | 0.0376510369543043 | 0.75709692 | 6.25150329395528e-90 |
| SC | ~~ | SC | 0.034079343 | 0.0017576229339658 | 0.47126817 | 0.0243053945839796 | 0.47126815 | 9.47439598214619e-84 |
| SI | ~~ | SI | 0.061614113 | 0.00251926594661925 | 0.43679668 | 0.0178596765734492 | 0.43679674 | 4.22192165152818e-132 |
| ALCDEP | ~~ | ALCDEP | 0.001093111 | 0.0135863316654366 | 0.01165573 | 0.145143683027888 | 0.01165596 | 0.935874051750451 |
| CUD | ~~ | CUD | 0.007519169 | 0.00301961512963333 | 0.12688524 | 0.0509561350905864 | 0.12688525 | 0.0127704195434713 |
| NICDEP | ~~ | NICDEP | 0.049337287 | 0.0110210887009626 | 0.57844472 | 0.129214129967346 | 0.57844392 | 7.58320921640698e-06 |
| OPIDEP | ~~ | OPIDEP | 0.001118851 | 0.0109504558915483 | 0.01136971 | 0.111334150918629 | 0.01136975 | 0.918618623417309 |
| TOWNSEND | ~~ | TOWNSEND | 0.010172334 | 0.00133901737748879 | 0.32376563 | 0.0426180865853358 | 0.32376520 | 3.03391728852635e-14 |
| TUD | ~~ | TUD | 0.014880273 | 0.00116843088221997 | 0.38560718 | 0.0302788200537068 | 0.38560759 | 3.76581524156498e-37 |
| EA | ~~ | EA | 0.015749256 | 0.00129066333471017 | 0.18669094 | 0.0152994806725004 | 0.18669097 | 3.01607838650667e-34 |
| AUDIT | ~~ | AUDIT | 0.084364976 | 0.00522506176139804 | 0.97665915 | 0.0604885913740527 | 0.97666167 | 1.20745057559347e-58 |
| CAN_USE | ~~ | CAN_USE | 0.096067681 | 0.00610247439570006 | 0.94364639 | 0.0599430035201337 | 0.94364743 | 7.74319022382343e-56 |
| PROB_OPI | ~~ | PROB_OPI | 0.007089614 | 0.00460466900658513 | 0.21119963 | 0.137163097126427 | 0.21119682 | 0.123643810508449 |
| DPW | ~~ | DPW | 0.041865943 | 0.0015199152852505 | 0.97929727 | 0.0355527372314146 | 0.97929724 | 5.09122785382196e-167 |
| DSM_NICDEP | ~~ | DSM_NICDEP | 0.013209399 | 0.0123221328721446 | 0.16150010 | 0.150661365597128 | 0.16150117 | 0.28371738526184 |
| INCOME_F | ~~ | INCOME_F | 0.022949813 | 0.00134266827544458 | 0.31437260 | 0.018392190650689 | 0.31437252 | 1.68231067253662e-65 |
| OCCUPATION | ~~ | OCCUPATION | 0.039362728 | 0.00342128940389647 | 0.26756611 | 0.0232559289134603 | 0.26756587 | 1.24155309423259e-30 |
| SMK | =~ | CPD | 1.000000000 | | 1.00000000 | | 0.49285199 | <NA> |
| SUB | =~ | CUD | 1.000000000 | | 1.00000000 | | 0.93440609 | <NA> |
| SES | =~ | EA | 1.000000000 | | 1.00000000 | | 0.90183648 | <NA> |
| GSMK | ~~ | GSMK | 1.000000000 | | 1.00000000 | | 1.00000000 | <NA> |
| GSUB | ~~ | GSUB | 1.000000000 | | 1.00000000 | | 1.00000000 | <NA> |
| GSES | ~~ | GSES | 1.000000000 | | 1.00000000 | | 1.00000000 | <NA> |
| GSES | ~~ | GSMK | 0.000000000 | | 0.00000000 | | 0.00000000 | <NA> |
| GSUB | ~~ | GSMK | 0.000000000 | | 0.00000000 | | 0.00000000 | <NA> |
| GSES | ~~ | GSUB | 0.000000000 | | 0.00000000 | | 0.00000000 | <NA> |
| SMK | ~~ | SMK | 0.000000000 | | 0.00000000 | | 0.00000000 | <NA> |
| SUB | ~~ | SUB | 0.000000000 | | 0.00000000 | | 0.00000000 | <NA> |
| SES | ~~ | SES | 0.000000000 | | 0.00000000 | | 0.00000000 | <NA> |
| SMK | ~~ | SUB | 0.000000000 | | 0.00000000 | | 0.00000000 | <NA> |
| SMK | ~~ | SES | 0.000000000 | | 0.00000000 | | 0.00000000 | <NA> |
| SUB | ~~ | SES | 0.000000000 | | 0.00000000 | | 0.00000000 | <NA> |

**Supplementary Table 2.** Full results of our proposed Genomic SEM hierarchical model. The nomenclature of the latent factors differs from the one presented in the manuscript. In this table, GSES=Common SMK/SU/SES. GSUB = Common SU. GSMK = Unique Smoking. SES = GSES. SUB = GSU. SMK = GSMK.

**Deviations from the preregistration**

| Preregistration | Final study | Reason |
|---|---|---|
| The project consists of 3 sets of analyses:<br>(1) Mapping of the latent factor architecture<br>(2) Estimating new genetic effects for all millions of loci for the identified latent factors<br>(3) Post-GWAS analyses on the smoking-specific instrument | The project consists of 4 sets of analyses:<br>(1) Latent factor architecture mapping<br>(2) Evaluation of the latent Unique Smoking Factor<br>(3) Genetic correlations between the latent smoking factor and a range of mental health outcomes<br>(4) Examining causal associations of Unique Smoking with mental health through MR. | Analyses 1-3 are similar to the ones described on the preregistration. The fourth analysis to check for causal effects was originally planned as a separate research project. Eventually, we combined both. |

| | | |
|---|---|---|
| By using the range of functions of Genomic SEM, we will perform both data driven and hypothesis driven analysis. | 'Next, we used a theory-driven hierarchical Genomic SEM approach, as previously described by…' | Given the complexity of the final model we wanted to fit, data-driven approaches could not cover the two-layered factor decomposition that we could achieve using theoretical models. Data-driven approaches were initially used to understand the data structure. Similarly, all first-layer factors were tested individually, but only the full factor is presented. |
| Cannabis frequency (upcoming) | DSM-based Nicotine Dependence – Johnson et al 2025<br><br>Problematic Opioid Prescription use – Sanchez-Roige 2021 | For those traits with more recent GWAS summary statistics available than those mentioned in the preregistration, recent ones were used.<br>In addition, cannabis frequency was not used, as the newest GWAS was not released by the time data analysis started.<br>DSM-based nicotine dependence and problematic opioid prescription use GWAS summary statistics were added to the model as they represent key additions to the smoking and substance use factors. |

**Supplementary Table 3.** Changes in the preregistration.

### References


Adams, M. J., Streit, F., Meng, X., Awasthi, S., Adey, B. N., Choi, K. W., Chundru, V. K., Coleman, J. R. I., Ferwerda, B., Foo, J. C., Gerring, Z. F., Giannakopoulou, O., Gupta, P., Hall, A. S. M., Harder, A., Howard, D. M., Hübel, C., Kwong, A. S. F., Levey, D. F.,…McIntosh, A. M. (2025). Trans-ancestry genome-wide study of depression identifies 697 associations implicating cell types and pharmacotherapies. *Cell*, *188*(3). https://doi.org/10.1016/j.cell.2024.12.002

Akimova, E. T., Wolfram, T., Ding, X., Tropf, F. C., Mills, M. C., Akimova, E. T., Wolfram, T., Ding, X., Tropf, F. C., & Mills, M. C. (2024). Polygenic prediction of occupational status GWAS elucidates genetic and environmental interplay in intergenerational transmission, careers and health in UK Biobank. *Nature Human Behaviour*, *9*(2). https://doi.org/10.1038/s41562-024-02076-3

Cornelis, M. C., Byrne, E. M., Esko, T., Nalls, M. A., Ganna, A., Paynter, N., Monda, K. L., Amin, N., Fischer, K., Renstrom, F., Ngwa, J. S., Huikari, V., Cavadino, A., Nolte, I. M., Teumer, A., Yu, K., Marques-Vidal, P., Rawal, R., Manichaikul, A.,…Chasman, D. I. (2014). Genome-wide meta-analysis identifies six novel loci associated with habitual coffee consumption. *Molecular psychiatry*, *20*(5). https://doi.org/10.1038/mp.2014.107

Gaddis, N., Mathur, R., Marks, J., Zhou, L., Quach, B., Waldrop, A., Levran, O., Agrawal, A., Randesi, M., Adelson, M., Jeffries, P. W., Martin, N. G., Degenhardt, L., Montgomery, G. W., Wetherill, L., Lai, D., Bucholz, K., Foroud, T., Porjesz, B.,…Johnson, E. O. (2022). Multi-trait genome-wide association study of opioid addiction: OPRM1 and beyond. *Scientific reports*, *12*(1). https://doi.org/10.1038/s41598-022-21003-y

Hatoum, A. S., Colbert, S. M. C., Johnson, E. C., Huggett, S. B., Deak, J. D., Pathak, G. A., Jennings, M. V., Paul, S. E., Karcher, N. R., Hansen, I., Baranger, D. A. A., Edwards, A., Grotzinger, A. D., Tucker-Drob, E. M., Kranzler, H. R., Davis, L. K., Sanchez-Roige, S., Polimanti, R., Gelernter, J.,…Agrawal, A. (2023). Multivariate genome-wide association meta-analysis of over 1 million subjects identifies loci underlying multiple substance use disorders. *Nature Mental Health*, *1*(3). https://doi.org/10.1038/s44220-023-00034-y

Jiang, L., Zheng, Z., Qi, T., Kemper, K. E., Wray, N. R., Visscher, P. M., & Yang, J. (2019). *A resource-efficient tool for mixed model association analysis of large-scale data* (1546-1718).

Johnson, E. C., Lai, D., Balbona, J. V., Miller, A. P., Hatoum, A. S., Deak, J. D., Jennings, M., Baranger, D. A. A., Galimberti, M., Sanichwankul, K., Thorgeirsson, T., Colbert, S. M. C., Adhikari, K., Docherty, A. R., Degenhardt, L., Edwards, T., Fox, L., Giannelis, A., Jeffries, P. W.,…Agrawal, A. (2025). Multi-ancestral genome-wide association study of clinically defined nicotine dependence reveals strong genetic correlations with other substance use disorders and health-related traits. *Psychological Medicine*, *55*. https://doi.org/10.1017/S0033291725100883

Kweon, H., Burik, C. A. P., Ning, Y., Ahlskog, R., Xia, C., Abner, E., Bao, Y., Bhatta, L., Faquih, T. O., de Feijter, M., Fisher, P., Gelemanović, A., Giannelis, A., Hottenga, J.-J., Khalili, B., Lee, Y., Li-Gao, R., Masso, J., Myhre, R.,…Koellinger, P. D. (2025). Associations between common genetic variants and income provide insights about the socio-economic health gradient. *Nature Human Behaviour*, *9*(4). https://doi.org/10.1038/s41562-024-02080-7

Levey, D. F., Galimberti, M., Deak, J. D., Wendt, F. R., Bhattacharya, A., Koller, D., Harrington, K. M., Quaden, R., Johnson, E. C., Gupta, P., Biradar, M., Lam, M., Cooke, M., Rajagopal, V. M., Empke, S. L. L., Zhou, H., Nunez, Y. Z., Kranzler, H. R., Edenberg, H. J.,…Gelernter, J. (2023). Multi-ancestry genome-wide association study of cannabis use disorder yields insight into disease biology and public health implications. *Nature genetics*, *55*(12). https://doi.org/10.1038/s41588-023-01563-z

Nievergelt, C. M., Maihofer, A. X., Atkinson, E. G., Chen, C.-Y., Choi, K. W., Coleman, J. R. I., Daskalakis, N. P., Duncan, L. E., Polimanti, R., Aaronson, C., Amstadter, A. B., Andersen, S. B., Andreassen,

O. A., Arbisi, P. A., Ashley-Koch, A. E., Austin, S. B., Avdibegoviç, E., Babić, D., Bacanu, S.-A.,…Koenen, K. C. (2024). Genome-wide association analyses identify 95 risk loci and provide insights into the neurobiology of post-traumatic stress disorder. *Nature genetics*, *56*(5). https://doi.org/10.1038/s41588-024-01707-9

O'Connell, K. S., Koromina, M., van der Veen, T., Boltz, T., David, F. S., Yang, J. M. K., Lin, K.-H., Wang, X., Coleman, J. R. I., Mitchell, B. L., McGrouther, C. C., Rangan, A. V., Lind, P. A., Koch, E., Harder, A., Parker, N., Bendl, J., Adorjan, K., Agerbo, E.,…Andreassen, O. A. (2025). Genomics yields biological and phenotypic insights into bipolar disorder. *Nature*, *639*(8056). https://doi.org/10.1038/s41586-024-08468-9

Okbay, A., Wu, Y., Wang, N., Jayashankar, H., Bennett, M., Nehzati, S. M., Sidorenko, J., Kweon, H., Goldman, G., Gjorgjieva, T., Jiang, Y., Hicks, B., Tian, C., Hinds, D. A., Ahlskog, R., Magnusson, P. K. E., Oskarsson, S., Hayward, C., Campbell, A.,…Young, A. I. (2022). Polygenic prediction of educational attainment within and between families from genome-wide association analyses in 3 million individuals. *Nature genetics*, *54*(4). https://doi.org/10.1038/s41588-022-01016-z

Pasman, J. A., Verweij, K. J. H., Gerring, Z., Stringer, S., Sanchez-Roige, S., Treur, J. L., Abdellaoui, A., Nivard, M. G., Baselmans, B. M. L., Ong, J.-S., Ip, H. F., van der Zee, M. D., Bartels, M., Day, F. R., Fontanillas, P., Elson, S. L., de Wit, H., Davis, L. K., MacKillop, J.,…Vink, J. M. (2018). GWAS of lifetime cannabis use reveals new risk loci, genetic overlap with psychiatric traits, and a causal effect of schizophrenia liability. *Nature Neuroscience*, *21*(9). https://doi.org/10.1038/s41593-018-0206-1

Quach, B. C., Bray, M. J., Gaddis, N. C., Liu, M., Palviainen, T., Minica, C. C., Zellers, S., Sherva, R., Aliev, F., Nothnagel, M., Young, K. A., Marks, J. A., Young, H., Carnes, M. U., Guo, Y., Waldrop, A., Sey, N. Y. A., Landi, M. T., McNeil, D. W.,…Hancock, D. B. (2020). Expanding the genetic architecture of nicotine dependence and its shared genetics with multiple traits. *Nature communications*, *11*(1). https://doi.org/10.1038/s41467-020-19265-z

Sanchez-Roige, S., Fontanillas, P., Jennings, M. V., Bianchi, S. B., Huang, Y., Hatoum, A. S., Sealock, J., Davis, L. K., Elson, S. L., Palmer, A. A., Sanchez-Roige, S., Fontanillas, P., Jennings, M. V., Bianchi, S. B., Huang, Y., Hatoum, A. S., Sealock, J., Davis, L. K., Elson, S. L., & Palmer, A. A. (2021). Genome-wide association study of problematic opioid prescription use in 132,113 23andMe research participants of European ancestry. *Molecular psychiatry*, *26*(11). https://doi.org/10.1038/s41380-021-01335-3

Sanchez-Roige, S., Palmer, A. A., Fontanillas, P., Elson, S. L., the 23andMe Research Team, t. S. U. D. W. G. o. t. P. G. C., Adams, M. J., Howard, D. M., Edenberg, H. J., Davies, G., Crist, R. C., Deary, I. J., McIntosh, A. M., & Clarke, T.-K. (2018). Genome-Wide Association Study Meta-Analysis of the Alcohol Use Disorders Identification Test (AUDIT) in Two Population-Based Cohorts. *American Journal of Psychiatry*, *176*(2). https://doi.org/10.1176/appi.ajp.2018.18040369

Saunders, G. R. B., Wang, X., Chen, F., Jang, S.-K., Liu, M., Wang, C., Gao, S., Jiang, Y., Khunsriraksakul, C., Otto, J. M., Addison, C., Akiyama, M., Albert, C. M., Aliev, F., Alonso, A., Arnett, D. K., Ashley-Koch, A. E., Ashrani, A. A., Barnes, K. C.,…Vrieze, S. (2022). Genetic diversity fuels gene discovery for tobacco and alcohol use. *Nature*, *612*(7941). https://doi.org/10.1038/s41586-022-05477-4

Strom, N. I., Gerring, Z. F., Galimberti, M., Yu, D., Halvorsen, M. W., Abdellaoui, A., Rodriguez-Fontenla, C., Sealock, J. M., Bigdeli, T., Coleman, J. R., Mahjani, B., Thorp, J. G., Bey, K., Burton, C. L., Luykx, J. J., Zai, G., Alemany, S., Andre, C., Askland, K. D.,…Mattheisen, M. (2025). Genome-wide analyses identify 30 loci associated with obsessive–compulsive disorder. *Nature genetics*, *57*(6). https://doi.org/10.1038/s41588-025-02189-z

Strom, N. I., Verhulst, B., Bacanu, S.-A., Cheesman, R., Purves, K. L., Gedik, H., Mitchell, B. L., Kwong, A. S., Faucon, A. B., Singh, K., Medland, S., Colodro-Conde, L., Krebs, K., Hoffmann, P., Herms, S., Gehlen, J., Ripke, S., Awasthi, S., Palviainen, T.,…Hettema, J. M. (2026). Genome-wide association study of major anxiety disorders in 122,341 European-ancestry cases identifies

58 loci and highlights GABAergic signaling. *Nature genetics*, *58*(2). https://doi.org/10.1038/s41588-025-02485-8

Toikumo, S., Jennings, M. V., Pham, B. K., Lee, H., Mallard, T. T., Bianchi, S. B., Meredith, J. J., Vilar-Ribó, L., Xu, H., Hatoum, A. S., Johnson, E. C., Pazdernik, V. K., Jinwala, Z., Pakala, S. R., Leger, B. S., Niarchou, M., Ehinmowo, M., Jenkins, G. D., Batzler, A.,…Sanchez-Roige, S. (2024). Multi-ancestry meta-analysis of tobacco use disorder identifies 461 potential risk genes and reveals associations with multiple health outcomes. *Nature Human Behaviour*, *8*(6). https://doi.org/10.1038/s41562-024-01851-6

Trubetskoy, V., Pardiñas, A. F., Qi, T., Panagiotaropoulou, G., Awasthi, S., Bigdeli, T. B., Bryois, J., Chen, C.-Y., Dennison, C. A., Hall, L. S., Lam, M., Watanabe, K., Frei, O., Ge, T., Harwood, J. C., Koopmans, F., Magnusson, S., Richards, A. L., Sidorenko, J.,…O'Donovan, M. C. (2022). Mapping genomic loci implicates genes and synaptic biology in schizophrenia. *Nature*, *604*(7906). https://doi.org/10.1038/s41586-022-04434-5

Walters, R. K., Polimanti, R., Johnson, E. C., McClintick, J. N., Adams, M. J., Adkins, A. E., Aliev, F., Bacanu, S.-A., Batzler, A., Bertelsen, S., Biernacka, J. M., Bigdeli, T. B., Chen, L.-S., Clarke, T.-K., Chou, Y.-L., Degenhardt, F., Docherty, A. R., Edwards, A. C., Fontanillas, P.,…Agrawal, A. (2018). Transancestral GWAS of alcohol dependence reveals common genetic underpinnings with psychiatric disorders. *Nature Neuroscience*, *21*(12). https://doi.org/10.1038/s41593-018-0275-1

Watanabe, K., Jansen, P. R., Savage, J. E., Nandakumar, P., Wang, X., Hinds, D. A., Gelernter, J., Levey, D. F., Polimanti, R., Stein, M. B., Van Someren, E. J. W., Smit, A. B., Posthuma, D., Watanabe, K., Jansen, P. R., Savage, J. E., Nandakumar, P., Wang, X., Hinds, D. A.,…Posthuma, D. (2022). Genome-wide meta-analysis of insomnia prioritizes genes associated with metabolic and psychiatric pathways. *Nature genetics*, *54*(8). https://doi.org/10.1038/s41588-022-01124-w

Watson, H. J., Yilmaz, Z., Thornton, L. M., Hübel, C., Coleman, J. R. I., Gaspar, H. A., Bryois, J., Hinney, A., Leppä, V. M., Mattheisen, M., Medland, S. E., Ripke, S., Yao, S., Giusti-Rodríguez, P., Hanscombe, K. B., Purves, K. L., Adan, R. A. H., Alfredsson, L., Ando, T.,…Bulik, C. M. (2019). Genome-wide association study identifies eight risk loci and implicates metabo-psychiatric origins for anorexia nervosa. *Nature genetics*, *51*(8). https://doi.org/10.1038/s41588-019-0439-2

Willer, C. J., Li, Y., & Abecasis, G. R. (2010). METAL: fast and efficient meta-analysis of genomewide association scans. *Bioinformatics*, *26*(17). https://doi.org/10.1093/bioinformatics/btq340
